# Association between household composition and severe COVID-19 outcomes in older people by ethnicity: an observational cohort study using the OpenSAFELY platform

**DOI:** 10.1101/2022.04.22.22274176

**Authors:** Kevin Wing, Daniel J Grint, Rohini Mathur, Hamish P Gibbs, George Hickman, Emily Nightingale, Anna Schultze, Harriet Forbes, Vahé Nafilyan, Krishnan Bhaskaran, Elizabeth Williamson, Thomas House, Lorenzo Pellis, Emily Herrett, Nileesa Gautam, Helen J Curtis, Christopher T Rentsch, Angel YS Wong, Brian MacKenna, Amir Mehrkar, Seb Bacon, Ian J Douglas, Stephen JW Evans, Laurie Tomlinson, Ben Goldacre, Rosalind M Eggo, the OpenSAFELY consortium

## Abstract

Ethnic differences in the risk of severe COVID-19 may be linked to household composition. We quantified the association between household composition and risk of severe COVID-19 by ethnicity for older individuals. With the approval of NHS England, we analysed ethnic differences in the association between household composition and severe COVID-19 in people aged 67 or over in England. We defined households by number of generations living together, and used multivariable Cox regression stratified by location and wave of the pandemic and accounted for age, sex, comorbidities, smoking, obesity, housing density and deprivation. We included 2 692 223 people over 67 years in wave 1 (01/02/2020-31/08/2020) and 2 731 427 in wave 2 (01/09/2020-31/01/2021). Multigenerational living was associated with increased risk of severe COVID-19 for White and South Asian older people in both waves (e.g. wave 2, 67+ living with 3 other generations vs 67+ year olds only: White HR 1·61 95% CI 1·38-1·87, South Asian HR 1·76 95% CI 1·48-2·10), with a trend for increased risks of severe COVID-19 with increasing generations in wave 2. Multigenerational living was associated with severe COVID-19 in older adults. Older South Asian people are over-represented within multigenerational households in England, especially in the most deprived settings. The number of generations in a household, number of occupants, ethnicity and deprivation status are important considerations in the continued roll-out of COVID-19 vaccination and targeting of interventions for future pandemics.

**Funding:** This research was funded in part, by the Wellcome Trust. For the purpose of open access, the author has applied a CC-BY public copyright licence to any Author Accepted Manuscript version arising from this submission.

## Introduction

The composition of a household - the age and number of its members - is a key determinant of infection risk for many infections, including COVID-19.^1–5^ Households with multiple generations may be at higher risk of infection due to increased routes of household introduction, with increased contact between older adults and working age adults of particular concern. Differences in the proportion of multigenerational households by ethnicity may be an underlying factor in the disproportionate effect that COVID-19 has had on ethnic minority groups in the UK.^1,6–11^ Analysing how multigenerational living affects the risk of severe COVID-19 in people of retirement age (over 67 in the UK) across wave 1 and wave 2 of the pandemic could improve our understanding of drivers of ethnic disparities in COVID-19 outcomes.

Using the OpenSAFELY platform, we sought to assess whether (1) household composition was associated with severe COVID-19 in older people within individual ethnic groups after accounting for potential confounders and (2) whether any association between household composition and severe COVID-19 and other potential household-level explanatory factors differed by ethnicity.

## Research in context

### Evidence before this study

We searched PubMed on 28 March 2022 for population-based studies examining the association between household composition and severe COVID-19 in older people. Keywords included (COVID-19 OR coronavirus OR SARS-CoV-2) AND (household) AND (mortality OR hospitalisation OR severe). We identified two studies, one from the UK and one from Sweden, both of which reported increased COVID-19 mortality in older people associated with multigenerational living. However, the study from Sweden was not able to assess whether results differed by ethnicity and did not account for comorbidities or investigate the role of other household-level variables (such as IMD or household size). The UK study did perform analysis by ethnicity but was based upon historical (2011 census) household composition data and did not examine effects separately by pandemic wave.

### Added value of this study

Our study is the first to use UK linked NHS primary and secondary electronic health records from February 2020 onwards to study the association between household composition and both hospitalisation and mortality due to COVID-19 in older people separately for wave 1 and wave 2 of the pandemic. Additionally, it is the only population-based study that uses up to date records to account for sociodemographic characteristics and clinical comorbidities while studying effects across ethnic groups, and is the first to allow the impact of increasing numbers of generations in a household on severe COVID-19 to be assessed. This has allowed us to demonstrate that living with more younger generations was associated with increased hazard of severe COVID-19 for White and South Asian older people in both waves, with a trend of increasing severe COVID-19 with increasing generations in wave 2. We were also able to demonstrate strong evidence that the effect of deprivation on severe COVID-19 was substantially greater for South Asian people compared to White people, an effect not seen for non-COVID outcomes, and that despite there being over ten times the number of older White people in England than older South Asian people, nearly twice the number of South Asian older people live in multigenerational houses in the most deprived settings. For older occupants of these households, rates of severe COVID-19 during wave 2 were higher than those for older people with multiple (established severe COVID-19 risk factor) comorbidities.

### Implications of all the available evidence

Multigenerational living is associated with severe COVID-19 in White and South Asian older people in England. Deprivation has a larger effect on severe COVID for South Asian people than White people, suggesting that disparities in COVID outcomes by ethnicity cannot be explained by socioeconomic factors alone. Older South Asian people are over-represented within multigenerational households in the most deprived settings, and experienced very high rates of severe COVID-19 during wave 2. Consideration of ethnicity, number of generations in a household (and/or total number of occupants in a household) and deprivation is warranted in planning booster vaccination roll-out and public health interventions (for future COVID-19 variants and other respiratory pandemic viruses).

## Methods

### Study design and population

We used linked primary care electronic health record data for 24 million people in England from the OpenSAFELY-TPP platform (see supplementary material). We extracted separate study populations for waves one (1 February 2020 to 31 August 2020) and two (1 September 2020 to 31 January 2021). We selected people aged 67 years or older at the start of each wave to represent a population (1) at or over retirement age and (2) at risk of severe COVID-19 due to their age. We wanted to assess if people of retirement age were at a differential risk of severe COVID-19 if they lived with younger generations who were more likely to be working (or attending educational or childcare settings). Participants were followed-up until the earliest of: the outcome of interest, deregistration from their general practice, death from any cause, or the end date for each wave. Our pre-specified study protocol (https://github.com/opensafely/hh-classification-research/tree/master/docs) and (post hoc) deviations from the protocol (supplementary table S1) are available.

#### Inclusion and exclusion criteria

Participants with at least 3 months of follow-up before the study start date for each wave were included, to ensure adequate capture of baseline factors. We used a TPP-developed pseudonymised household identifier which links people living at the same address on 1 February 2020,^12^ (see supplementary methods). We excluded people with no household identifier, households with anyone flagged as living in a care home (with care home status derived by matching addresses to Care Quality Commission data^13^), those living with more than 12 people (possible care homes or other institutions), and those living with more than 4 people who were all over the age of 67 (possible care homes).^13^ We also excluded people with missing sex, location (based on Middle Layer Super Output Area - MSOA) or index of multiple deprivation (indicators of poor data quality when missing).

### Exposure

We classified distinct generations as 0-17 year olds, 18-29 year olds, 30-66 year olds and 67+ year olds before assigning each 67+ year old in our study to one of the following five household composition categories:

**67+ living alone:** 67+ year old living alone (single occupancy household)

**Multiple 67+ year olds:** 67+ year old living with up to three other 67+ year olds (reference category)

**1 other generation**: 67+ year old(s) living with people from just ONE other younger generation

**2 other generations:** 67+ year old(s) living with people from TWO other younger generations

**3 other generations:** 67+ year old(s) living with people from all THREE other younger generations

The primary exposure was the household composition where each 67+ year old was resident on 1st February 2020.

### Outcomes

The primary outcome was “severe COVID-19” defined as COVID-19 related hospital admission or death between 1 February 2020 and 31 August 2020 (for wave 1), and 1

September 2020 and 31 January 2021 (for wave 2):

1. Hospital admission with COVID-19: a COVID-19 ICD-10 code for confirmed (U07.1) or suspected (U07.2) COVID-19 in the primary diagnosis field in Secondary Use Service (SUS) data
2. COVID-19-related death: a COVID-19 ICD-10 code for confirmed or suspected COVID-19 anywhere on the death certificate.

We also analysed each of the above outcomes separately, and included a secondary outcome of “non-COVID-19 death” (death from any other cause on the death certificate) to assess specificity of results.

### Covariates

#### Ethnicity

We investigated the effect of household composition within GP-recorded census ethnicity categories of White, South Asian, Black, mixed and other. Detailed census categories (Table S2) were analysed for any ethnicity where household composition was associated with severe COVID-19.

#### Other covariates

We included categorical age in years (67-69, 70-74, 75-79, 80-84, 85+), sex, body mass index categories (kg/m^2^) (underweight, normal, overweight, obese I, obese II, obese III), index of multiple deprivation quintiles (from 1 - most affluent to 5 - most deprived), geographic region defined by Upper Tier Local Authority (UTLA), smoking status (current, former, never), housing density (ONS Rural/Urban classification in categories of urban major/minor conurbation, urban city and town, rural town, rural village) and number of comorbidities shown to be associated with poor COVID-19 outcomes (0, 1 or 2+)^14^ (see Supplementary Material Tables S3 and S4).

### Statistical analysis

#### Descriptive analysis

Participant characteristics at baseline were summarised by household composition category separately for wave 1 and for wave 2.

#### Household composition and severe COVID-19/non-COVID-19 death by ethnicity

We used multivariable Cox proportional hazards regression with robust standard errors to account for clustering by household in order to estimate differences by household composition in the hazard of severe COVID-19 and non-COVID-19 death. All models were stratified by UTLA region to account for region-specific variation in infection rates over time,^15^ and separate analyses were performed for wave 1 and wave 2. An interaction between household composition and ethnicity was included in all models, with all household composition results reported by ethnicity. We hypothesised that associations between other covariates and severe COVID-19 would vary by ethnicity, so performed likelihood ratio tests for interaction (LRT) between each covariate and ethnicity (including interactions as appropriate in our final models). Where deprivation or housing density interacted with ethnicity, we also present results stratified by ethnicity for these variables.

We assessed the proportional hazards assumption by testing for a zero slope in the scaled Schoenfeld residuals and through graphical inspection of plots of the Schoenfeld residuals against time.

#### Analysis of the impact of household size (number of occupants)

In our main analysis of household composition we decided a-priori not to adjust for household size (see supplementary methods). Instead, to assess how the hazard of severe COVID varied by number of occupants within categories of household composition and vice versa, we cross-tabulated results from a combined household composition-household size exposure variable (Supplementary Table S6).

#### Absolute rates of severe COVID-19 and non-COVID-19 death by household characteristics

Rates of all outcomes were reported by ethnicity and (1) household composition (2) household size (3) any other household-level explanatory variables found to differ by ethnicity and (4) those with two or more comorbidities (for comparison).

#### Sensitivity analyses

We tested the impact of including a 5 year “buffer” between the 67+ year old generation and the next youngest generation (to avoid a 67+ year old living with a 61-66 year old being considered multigenerational). We assessed the impact of only including people who lived in households with 100% TPP coverage (i.e. all adults in the household were registered with TPP - see supplementary methods). Our main analysis was a complete ethnicity records analysis - we performed a sensitivity analysis applying multiple imputation to account for missing ethnicity (10 imputations). In our main analysis people with missing body mass index were assumed to be normal weight, and those with missing smoking data were assumed to be never-smokers (on the assumption that obesity and smoking would likely be recorded if present) - a complete records sensitivity analysis for BMI and smoking was performed.

### Software and Reproducibility

This analysis was delivered through the OpenSAFELY platform - see supplementary material for details.

### Patient and Public Involvement

We have developed a publicly available website https://opensafely.org/ through which we invite any patient or member of the public to contact us regarding this study or the broader OpenSAFELY project.

## Results

### Descriptive results

From a total of 23 696 832 individuals in the OpenSAFELY database on 1 February, 2020, there were 2 692 223 people aged 67 or over (referred to as: “67+”) at the beginning of wave 1 who met the selection criteria (Figure S1). Of these 1 109 443 (41·2%) lived with other 67+, 920 670 (34·2%) lived alone, 526 037 (19·5%) lived with one other younger generation, 113 553 (4·2%) lived with two other younger generations and 22 540 (0·8%) lived with three other younger generations (Table 1). The final wave 2 cohort was slightly larger (2 731 427 people), with no change in relative proportion of household compositions (Table 1).

**Table 1:** Baseline characteristics of cohort of 67+ year olds, by categories of generational household composition during the first and second waves of the pandemic in the UK.

| Characteristic<sup>1</sup> | Wave 1 (1<sup>st</sup> February – 31<sup>st</sup> August 2020) |  |  |  |  |  | Wave 2 (1<sup>st</sup> September – 31<sup>st</sup> January 2021) |  |  |  |  |  |
| --- | --- | --- | --- | --- | --- | --- | --- | --- | --- | --- | --- | --- |
|  | Total<br>(n=2692223) | Household composition (number of generations in household) |  |  |  |  | Total<br>(n=2731427) | Household composition (number of generations in household) |  |  |  |  |
|  |  | Multiple 67+<br>year olds<br>(n=1109443) | 67+ living alone<br>(n=920670) | 67+ & 1<br>(n=526037) | 67+ & 2<br>(n=113533) | 67+ & 3<br>(n=22540) |  | Multiple 67+<br>year olds<br>(n=1143558) | 67+ living alone<br>(n=914039) | 67+ & 1<br>(n=534276) | 67+ & 2<br>(n=116278) | 67+ & 3<br>(n=23276) |
| Sex |  |  |  |  |  |  |  |  |  |  |  |  |
| F | 1450088 (53.9) | 560567 (50.5) | 586899 (63.7) | 237493 (45.1) | 53200 (46.9) | 11929 (52.9) | 1470941 (53.9) | 580492 (50.8) | 581276 (63.6) | 242588 (45.4) | 54212 (46.6) | 12373 (53.2) |
| M | 1242135 (46.1) | 548876 (49.5) | 333771 (36.3) | 288544 (54.9) | 60333 (53.1) | 10611 (47.1) | 1260486 (46.1) | 563066 (49.2) | 332763 (36.4) | 291688 (54.6) | 62066 (53.4) | 10903 (46.8) |
| Age (years) |  |  |  |  |  |  |  |  |  |  |  |  |
| Mean (SD) | 75.8 (6.7) | 75.5 (5.8) | 77.8 (7.5) | 73.5 (6.2) | 73.7 (6.2) | 74.5 (6.5) | 75.9 (6.7) | 75.6 (5.9) | 77.9 (7.5) | 73.6 (6.2) | 73.6 (6.2) | 74.5 (6.5) |
| Median (IQR) | 74.0 (70.0-80.0) | 74.0 (71.0-79.0) | 77.0 (72.0-83.0) | 72.0 (69.0-77.0) | 72.0 (69.0-77.0) | 73.0 (69.0-79.0) | 74.0 (71.0-80.0) | 75.0 (71.0-79.0) | 77.0 (72.0-83.0) | 72.0 (69.0-77.0) | 72.0 (69.0-77.0) | 73.0 (69.0-79.0) |
| Categorical |  |  |  |  |  |  |  |  |  |  |  |  |
| 67-69 | 503732 (18.7) | 163996 (14.8) | 128739 (14.0) | 168840 (32.1) | 35857 (31.6) | 6300 (28.0) | 505939 (18.5) | 164493 (14.4) | 127802 (14.0) | 170305 (31.9) | 36954 (31.8) | 6385 (27.4) |
| 70-74 | 856665 (31.8) | 393791 (35.5) | 237702 (25.8) | 180272 (34.3) | 37902 (33.4) | 6998 (31.0) | 869008 (31.8) | 402170 (35.2) | 236997 (25.9) | 183442 (34.3) | 39056 (33.6) | 7343 (31.5) |
| 75-79 | 592679 (22.0) | 286581 (25.8) | 194667 (21.1) | 87617 (16.7) | 19582 (17.2) | 4232 (18.8) | 605755 (22.2) | 297690 (26.0) | 193972 (21.2) | 89779 (16.8) | 19880 (17.1) | 4434 (19.0) |
| 80-84 | 403923 (15.0) | 170999 (15.4) | 167516 (18.2) | 50654 (9.6) | 11820 (10.4) | 2934 (13.0) | 408023 (14.9) | 177839 (15.6) | 163864 (17.9) | 51432 (9.6) | 11875 (10.2) | 3013 (12.9) |
| 85+ | 335224 (12.5) | 94076 (8.5) | 192046 (20.9) | 38654 (7.3) | 8372 (7.4) | 2076 (9.2) | 342702 (12.5) | 101366 (8.9) | 191404 (20.9) | 39318 (7.4) | 8513 (7.3) | 2101 (9.0) |
| Ethnicity |  |  |  |  |  |  |  |  |  |  |  |  |
| White | 2548262 (94.7) | 1083552 (97.7) | 887343 (96.4) | 482696 (91.8) | 82362 (72.5) | 12309 (54.6) | 2583394 (94.6) | 1116533 (97.6) | 880434 (96.3) | 489303 (91.6) | 84368 (72.6) | 12756 (54.8) |
| South Asian | 87132 (3.2) | 15264 (1.4) | 14674 (1.6) | 26557 (5.0) | 22650 (20.0) | 7987 (35.4) | 89653 (3.3) | 15969 (1.4) | 14745 (1.6) | 27605 (5.2) | 23136 (19.9) | 8198 (35.2) |
| Black | 26478 (1.0) | 4019 (0.4) | 9399 (1.0) | 7888 (1.5) | 4017 (3.5) | 1155 (5.1) | 26948 (1.0) | 4132 (0.4) | 9462 (1.0) | 8050 (1.5) | 4119 (3.5) | 1185 (5.1) |
| Mixed | 9394 (0.3) | 2027 (0.2) | 3129 (0.3) | 2711 (0.5) | 1209 (1.1) | 318 (1.4) | 9696 (0.4) | 2117 (0.2) | 3188 (0.3) | 2832 (0.5) | 1234 (1.1) | 325 (1.4) |
| Other | 20957 (0.8) | 4581 (0.4) | 6125 (0.7) | 6185 (1.2) | 3295 (2.9) | 771 (3.4) | 21736 (0.8) | 4807 (0.4) | 6210 (0.7) | 6486 (1.2) | 3421 (2.9) | 812 (3.5) |
| BMI<sup>2</sup> category |  |  |  |  |  |  |  |  |  |  |  |  |
| Underweight | 58508 (2.2) | 19078 (1.7) | 28043 (3.0) | 9027 (1.7) | 1899 (1.7) | 461 (2.0) | 32136 (1.2) | 11011 (1.0) | 15141 (1.7) | 4801 (0.9) | 978 (0.8) | 205 (0.9) |
| Normal<sup>3</sup> | 956951 (35.5) | 395450 (35.6) | 350630 (38.1) | 169195 (32.2) | 35184 (31.0) | 6492 (28.8) | 1784671 (65.3) | 748250 (65.4) | 609405 (66.7) | 338803 (63.4) | 73784 (63.5) | 14429 (62.0) |
| Overweight | 979424 (36.4) | 424681 (38.3) | 310771 (33.8) | 195590 (37.2) | 40785 (35.9) | 7597 (33.7) | 499221 (18.3) | 220083 (19.2) | 155953 (17.1) | 98704 (18.5) | 20539 (17.7) | 3942 (16.9) |
| Obese I | 478907 (17.8) | 191553 (17.3) | 155410 (16.9) | 102917 (19.6) | 23784 (20.9) | 5243 (23.3) | 276361 (10.1) | 112692 (9.9) | 86992 (9.5) | 60007 (11.2) | 13650 (11.7) | 3020 (13.0) |
| Obese II | 157352 (5.8) | 57988 (5.2) | 53931 (5.9) | 35082 (6.7) | 8404 (7.4) | 1947 (8.6) | 98799 (3.6) | 37341 (3.3) | 32776 (3.6) | 22375 (4.2) | 5112 (4.4) | 1195 (5.1) |
| Obese III | 61081 (2.3) | 20693 (1.9) | 21885 (2.4) | 14226 (2.7) | 3477 (3.1) | 800 (3.5) | 40239 (1.5) | 14181 (1.2) | 13772 (1.5) | 9586 (1.8) | 2215 (1.9) | 485 (2.1) |
| Smoking status |  |  |  |  |  |  |  |  |  |  |  |  |
| Never<sup>4</sup> | 1070550 (39.8) | 443554 (40.0) | 358348 (38.9) | 204251 (38.8) | 52581 (46.3) | 11816 (52.4) | 1089415 (39.9) | 458986 (40.1) | 355800 (38.9) | 208636 (39.1) | 53816 (46.3) | 12177 (52.3) |
| Former | 1408300 (52.3) | 605090 (54.5) | 473653 (51.4) | 271542 (51.6) | 49669 (43.7) | 8346 (37.0) | 1427167 (52.2) | 622275 (54.4) | 470381 (51.5) | 275153 (51.5) | 50746 (43.6) | 8612 (37.0) |
| Current | 213373 (7.9) | 60799 (5.5) | 88669 (9.6) | 50244 (9.6) | 11283 (9.9) | 2378 (10.6) | 214845 (7.9) | 62297 (5.4) | 87858 (9.6) | 50487 (9.4) | 11716 (10.1) | 2487 (10.7) |
| Index of multiple deprivation |  |  |  |  |  |  |  |  |  |  |  |  |
| 1 (affluent) | 664384 (24.7) | 324311 (29.2) | 196213 (21.3) | 118676 (22.6) | 22151 (19.5) | 3033 (13.5) | 676016 (24.7) | 334864 (29.3) | 195057 (21.3) | 120245 (22.5) | 22681 (19.5) | 3169 (13.6) |
| 2 | 625275 (23.2) | 280387 (25.3) | 200876 (21.8) | 117551 (22.3) | 22943 (20.2) | 3518 (15.6) | 645354 (23.6) | 293400 (25.7) | 203112 (22.2) | 121387 (22.7) | 23817 (20.5) | 3638 (15.6) |
| 3 | 577223 (21.4) | 237830 (21.4) | 196871 (21.4) | 113728 (21.6) | 24351 (21.4) | 4443 (19.7) | 581409 (21.3) | 242636 (21.2) | 194530 (21.3) | 114770 (21.5) | 24828 (21.4) | 4645 (20.0) |
| 4 | 477195 (17.7) | 167960 (15.1) | 179416 (19.5) | 99825 (19.0) | 24248 (21.4) | 5746 (25.5) | 476261 (17.4) | 170310 (14.9) | 175362 (19.2) | 100189 (18.8) | 24573 (21.1) | 5827 (25.0) |
|  | Total<br>(n=2692223) | Household composition (number of generations in household) |  |  |  |  | Total<br>(n=2731427) | Household composition (number of generations in household) |  |  |  |  |
|  |  | Multiple 67+<br>year olds<br>(n=1109443) | 67+ living alone<br>(n=920670) | 67+ & 1<br>(n=526037) | 67+ & 2<br>(n=113533) | 67+ & 3<br>(n=22540) |  | Multiple 67+<br>year olds<br>(n=1143558) | 67+ living alone<br>(n=914039) | 67+ & 1<br>(n=534276) | 67+ & 2<br>(n=116278) | 67+ & 3<br>(n=23276) |
| 5 (deprived) | 336834 (12·5) | 94155 (8·5) | 142899 (15·5) | 74609 (14·2) | 19424 (17·1) | 5747 (25·5) | 338945 (12·4) | 96509 (8·4) | 140941 (15·4) | 75708 (14·2) | 19865 (17·1) | 5922 (25·4) |
| Region of England |  |  |  |  |  |  |  |  |  |  |  |  |
| East | 631645 (23·5) | 270459 (24·4) | 205218 (22·3) | 124028 (23·6) | 26875 (23·7) | 5065 (22·5) | 641327 (23·5) | 278664 (24·4) | 203580 (22·3) | 126120 (23·6) | 27698 (23·8) | 5265 (22·6) |
| East Midlands | 499640 (18·6) | 215911 (19·5) | 162124 (17·6) | 97962 (18·6) | 19742 (17·4) | 3901 (17·3) | 506526 (18·5) | 222541 (19·5) | 160463 (17·6) | 99207 (18·6) | 20271 (17·4) | 4044 (17·4) |
| London | 119412 (4·4) | 23076 (2·1) | 43479 (4·7) | 34018 (6·5) | 15216 (13·4) | 3623 (16·1) | 120579 (4·4) | 23695 (2·1) | 43151 (4·7) | 34762 (6·5) | 15346 (13·2) | 3625 (15·6) |
| North East | 139434 (5·2) | 59213 (5·3) | 48600 (5·3) | 26450 (5·0) | 4361 (3·8) | 810 (3·6) | 140938 (5·2) | 60798 (5·3) | 48048 (5·3) | 26749 (5·0) | 4499 (3·9) | 844 (3·6) |
| North West | 262220 (9·7) | 108562 (9·8) | 93956 (10·2) | 50864 (9·7) | 7622 (6·7) | 1216 (5·4) | 265738 (9·7) | 111821 (9·8) | 93244 (10·2) | 51585 (9·7) | 7778 (6·7) | 1310 (5·6) |
| South East | 180063 (6·7) | 75065 (6·8) | 63875 (6·9) | 33450 (6·4) | 6668 (5·9) | 1005 (4·5) | 183145 (6·7) | 77631 (6·8) | 63667 (7·0) | 33945 (6·4) | 6846 (5·9) | 1056 (4·5) |
| South West | 408689 (15·2) | 181212 (16·3) | 139861 (15·2) | 72401 (13·8) | 13377 (11·8) | 1838 (8·2) | 416226 (15·2) | 187337 (16·4) | 139798 (15·3) | 73582 (13·8) | 13612 (11·7) | 1897 (8·2) |
| West Midlands | 94762 (3·5) | 32525 (2·9) | 34180 (3·7) | 20904 (4·0) | 5675 (5·0) | 1478 (6·6) | 95434 (3·5) | 33244 (2·9) | 33661 (3·7) | 21203 (4·0) | 5787 (5·0) | 1539 (6·6) |
| Yorkshire | 355818 (13·2) | 143251 (12·9) | 129154 (14·0) | 65847 (12·5) | 13964 (12·3) | 3602 (16·0) | 360972 (13·2) | 147644 (12·9) | 128220 (14·0) | 67005 (12·5) | 14409 (12·4) | 3694 (15·9) |
| Housing density |  |  |  |  |  |  |  |  |  |  |  |  |
| Conurbation |  |  |  |  |  |  |  |  |  |  |  |  |
| Urban major | 421379 (15·7) | 130132 (11·7) | 154613 (16·8) | 96556 (18·4) | 31749 (28·0) | 8329 (37·0) | 425489 (15·6) | 133554 (11·7) | 152870 (16·7) | 98309 (18·4) | 32266 (27·7) | 8490 (36·5) |
| Urban minor | 172486 (6·4) | 69736 (6·3) | 62353 (6·8) | 33533 (6·4) | 5792 (5·1) | 1072 (4·8) | 174064 (6·4) | 71620 (6·3) | 61529 (6·7) | 33850 (6·3) | 5927 (5·1) | 1138 (4·9) |
| Urban city & town | 1350131 (50·1) | 557061 (50·2) | 470702 (51·1) | 259693 (49·4) | 52802 (46·5) | 9873 (43·8) | 1368303 (50·1) | 573239 (50·1) | 466797 (51·1) | 263678 (49·4) | 54318 (46·7) | 10271 (44·1) |
| Rural |  |  |  |  |  |  |  |  |  |  |  |  |
| Town | 408668 (15·2) | 190703 (17·2) | 133360 (14·5) | 71820 (13·7) | 11232 (9·9) | 1553 (6·9) | 414986 (15·2) | 196876 (17·2) | 132392 (14·5) | 72628 (13·6) | 11496 (9·9) | 1594 (6·8) |
| Village | 328247 (12·2) | 157011 (14·2) | 95247 (10·3) | 62787 (11·9) | 11542 (10·2) | 1660 (7·4) | 335143 (12·3) | 162430 (14·2) | 95414 (10·4) | 63834 (11·9) | 11757 (10·1) | 1708 (7·3) |
| Number of comorbidities |  |  |  |  |  |  |  |  |  |  |  |  |
| 0 | 1087692 (40·4) | 480966 (43·4) | 331798 (36·0) | 222583 (42·3) | 44714 (39·4) | 7631 (33·9) | 1134668 (41·5) | 507918 (44·4) | 339683 (37·2) | 231972 (43·4) | 47004 (40·4) | 8091 (34·8) |
| 1 | 847951 (31·5) | 345372 (31·1) | 293795 (31·9) | 165060 (31·4) | 36367 (32·0) | 7357 (32·6) | 858490 (31·4) | 354537 (31·0) | 291709 (31·9) | 167136 (31·3) | 37432 (32·2) | 7676 (33·0) |
| 2+ | 756580 (28·1) | 283105 (25·5) | 295077 (32·1) | 138394 (26·3) | 32452 (28·6) | 7552 (33·5) | 738269 (27·0) | 281103 (24·6) | 282647 (30·9) | 135168 (25·3) | 31842 (27·4) | 7509 (32·3) |

Note 1: Data are n(%) unless specified. Note 2: BMI=Body Mass Index. Note 3: BMI “normal”includes those with missing data. Note 4: Smoking status “never” includes those with missing data.

London had the highest proportion of 67+ living in multigenerational houses (15·8%), with a much lower proportion in the North West (3·4%) (Figure 1). Younger 67+ and those living in urban areas were more likely to live with multiple younger generations (Table 1). A notably larger proportion of South Asian (66%) and Black people (49%) 67+ were living with one or more other generations than White 67+ (23%) (Figure 1). Household composition by ethnicity for wave 2 was similar to wave 1 (Supplementary Table S7).

**Figure 1.**
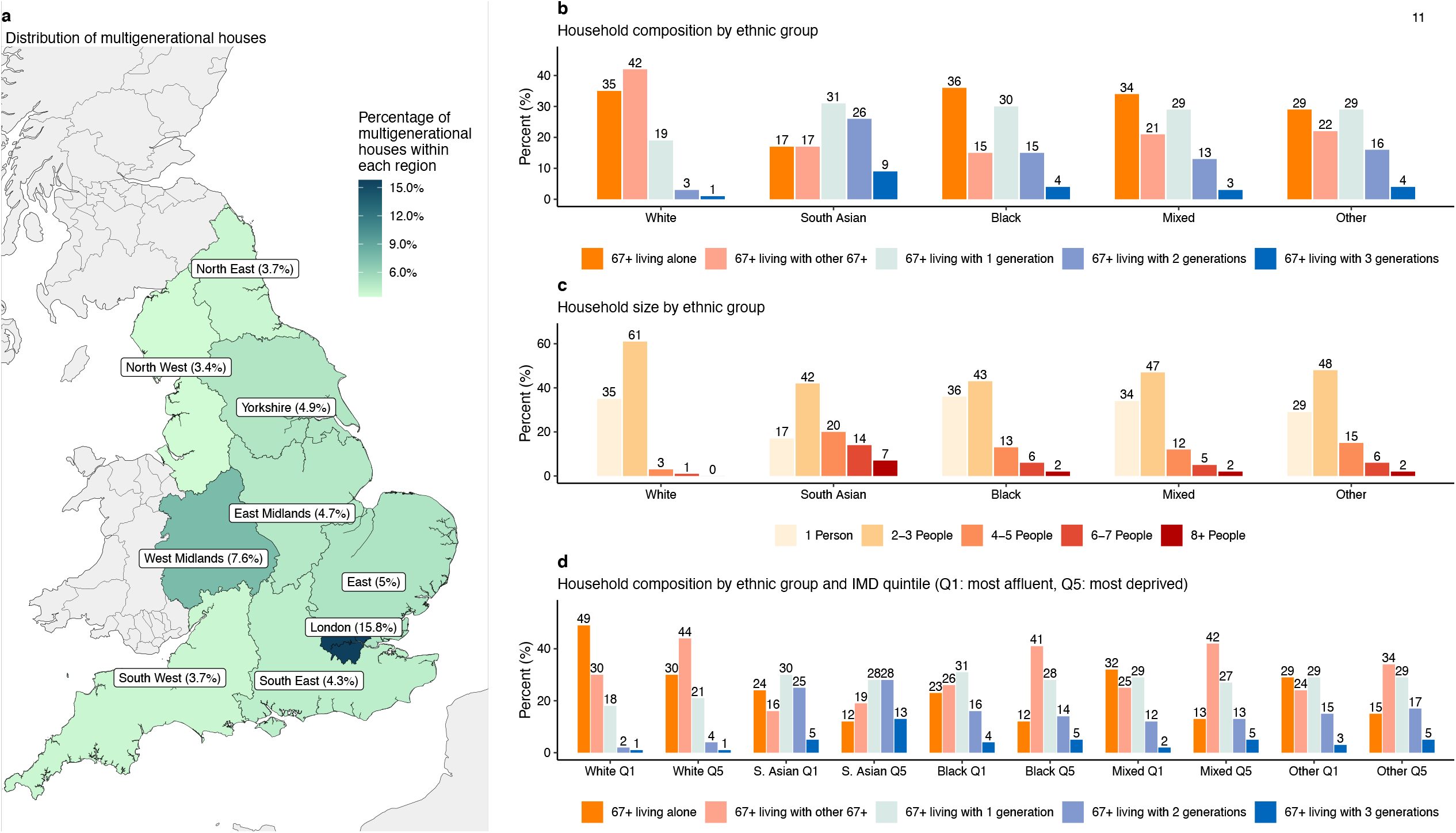
Summary population characteristics for households by ethnicity. *a*, Distribution of multigenerational houses by region of England; *b*, age-generational household composition by ethnic group; *c*, household size (total number of occupants) by ethnic group; *d*, age-generational household composition by ethnic group and IMD quintile (Q1: most affluent, Q5: most deprived). Abbreviations: IMD, index of multiple deprivation.

### Household composition and severe COVID-19 by ethnicity

#### Wave 1

After accounting for age, sex, comorbidities, housing density, deprivation status, obesity and smoking, and including interactions between ethnicity and household composition and age (Table S8), White 67+ living alone had an increased risk of severe COVID-19 compared to the reference group (HR 1·35 95% CI 1·30-1·41) (Figure 2 (W1)). There was a small increase in hazard for 67+ living with any other younger generation (e.g. 67+ 2 other generations: HR 1·22 95% CI 1·10-1·35) (Figure 2 (W1). For South Asian 67+, there was also an increase in hazard of severe COVID-19 in those living alone (HR 1·47 95% CI 1·18-1·84), and living with either 2 or 3 generations (e.g. 3 other generations HR 1·41 95% CI 1·09-1·83). For all other ethnicities wide confidence intervals limited interpretation, but estimates were generally consistent with an increased HR for multigenerational living (Figure 2 (W1).

**Figure 2.**
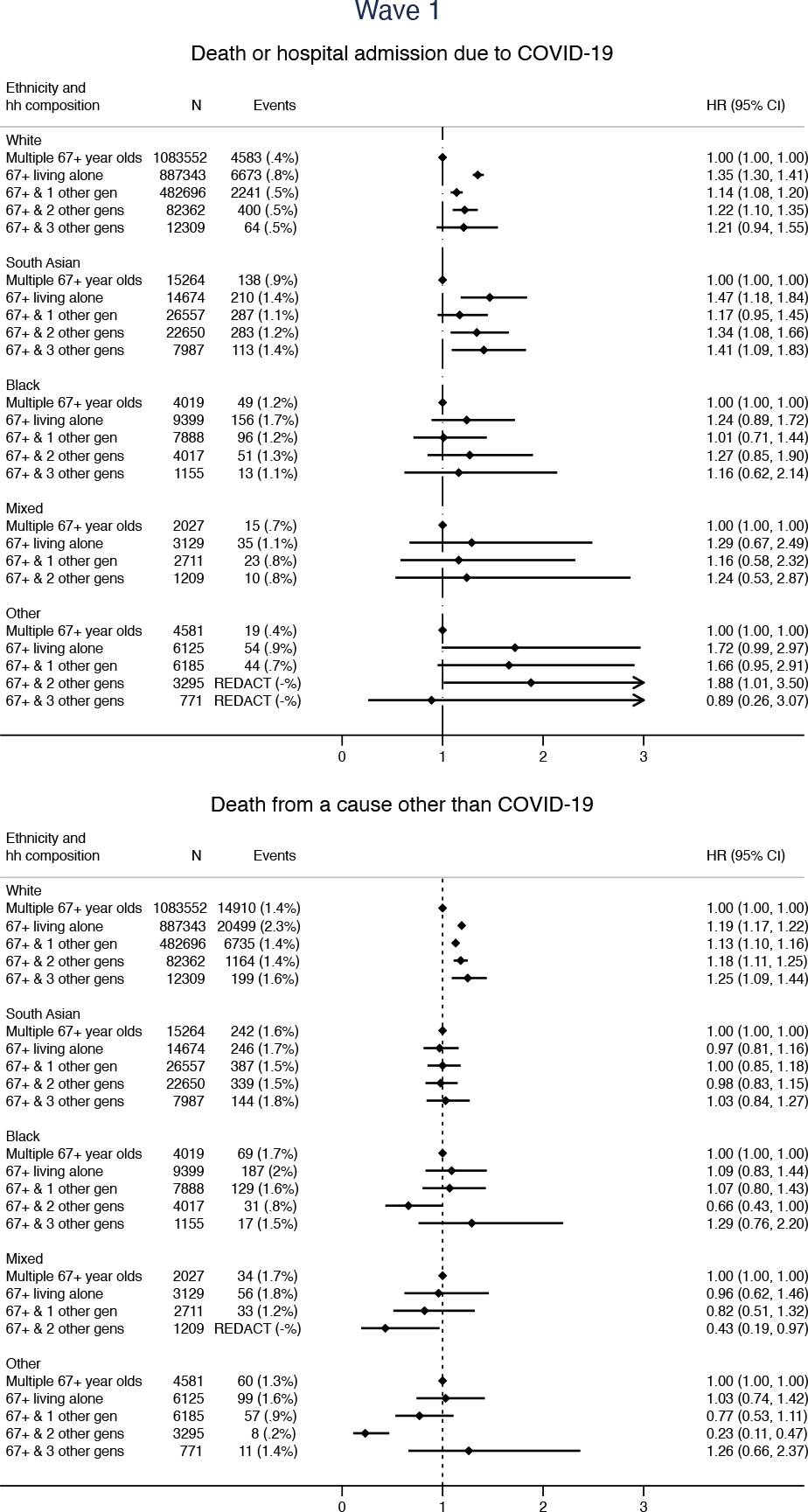
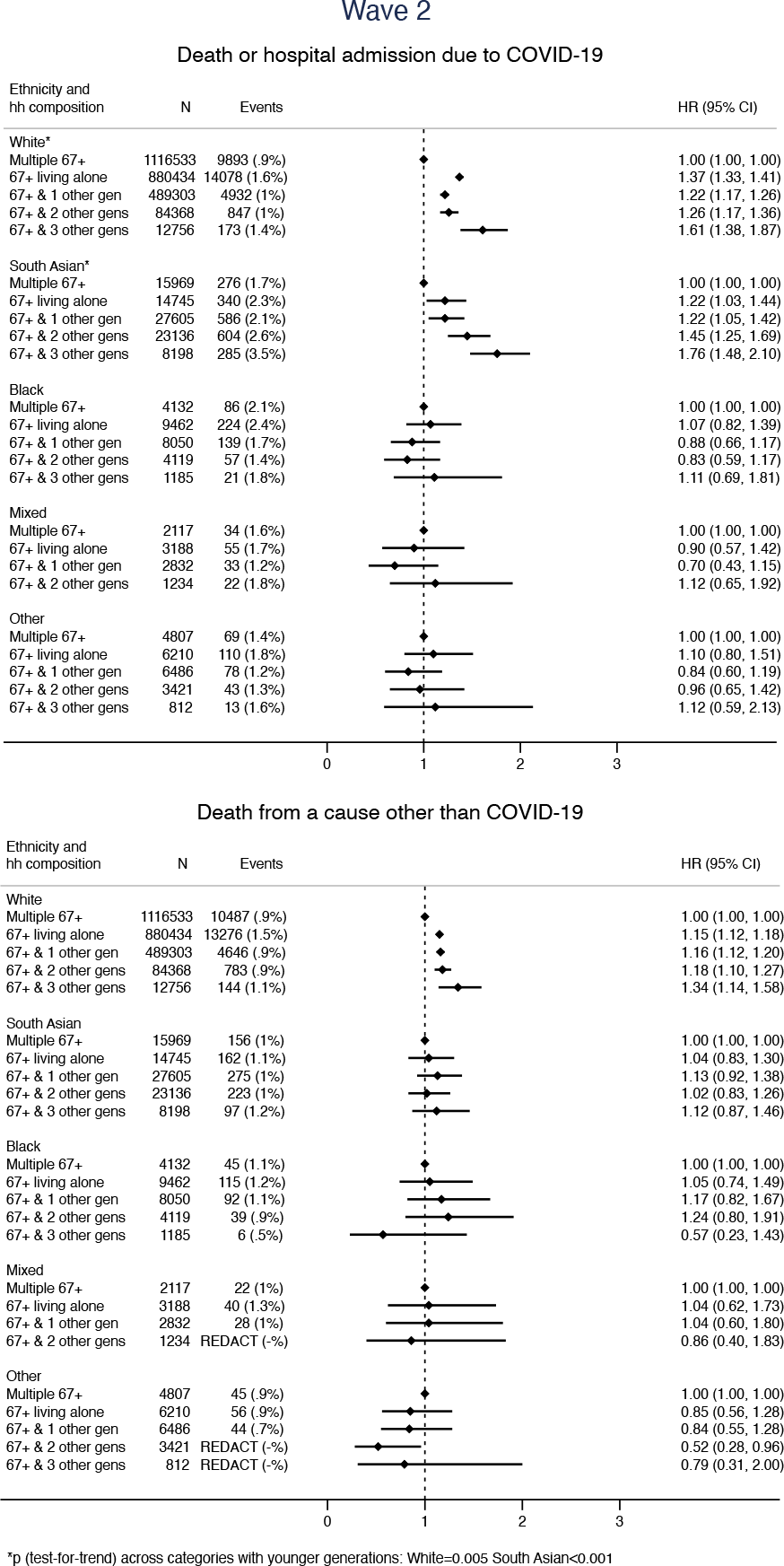
Association between household composition and (1) severe COVID-19 (death or hospitalisation due to COVID-19) and (2) non-COVID-19 death by ethnicity for wave 1 and wave 2 of the pandemic in England. Household composition in terms of number of distinct generations that each 67+ year old in the study cohort is living with (considering the following distinct generations: 0-17 year olds, 18-29 year olds, 30-66 year olds, 67+ year olds) i.e. 67+ & 1=67+ year old’s household includes one other younger generation, 67+ & 2=67+ year old’s household includes two other younger generations, 67+ & 3=67+ year old’s household includes three other younger generations. Wave 1 models stratified by location (UTLA) and adjusted for sex, number of comorbidities, categories of housing density (rural or urban setting), smoking status, socio-economic status and including an interaction between ethnicity and age (as well as the interaction between household composition and ethnicity presented here). Wave 2 models stratified on UTLA and adjusted for: sex, smoking, housing density and number of comorbidities and including interactions between ethnicity and: IMD, age and obesity (as well as the interaction with household composition presented here).

For White people, the association of household composition with non-COVID-19 death was similar to the association with severe COVID-19 while for South Asian people associations were specific to COVID-19 (e.g. non-COVID-19 death 67+ & 2 other generations: HR 0·98, 0·83-1·15 (Figure 2 (W1)). Despite wide confidence intervals, results for other ethnicities were consistent with those for South Asian people.

#### Wave 2

After accounting for sex, comorbidities, housing density and smoking, and including interactions between ethnicity and: household composition, age, deprivation status and obesity (Table S8), multigenerational living for White people in wave 2 was associated with higher hazards of severe COVID-19 compared to wave 1 (e.g. 67+ & 3 other generations: wave 2 HR 1·61 95% CI 1·38-1·87) (Figure 2 (W2)) and there was evidence for an increasing hazard of severe COVID-19 with increasing number of generations (Figure 2 (W2)). For South Asian people, a similar trend was observed (Figure 2 (W2)), with increases in HRs for all categories of multigenerational living compared to wave 1 (e.g. 67+ & 3 other generations wave 2 HR 1·76 95% CI 1·48-2·10) (Figure 2 (W2)).

For South Asian people, the specificity of effect persisted and increased for Wave 2 (e.g. Wave 2, 67+ & 3 other generations: severe COVID-19 HR 1·76 95% CI 1·48-2·10, non-COVID-19 death HR 1·12 95% CI 0·87-1·46) (Figure 2 (W2)). For Black and Mixed ethnicities, although wide confidence intervals limited interpretation, increased multigenerational living seemed to be associated with an increased hazard of non-COVID-19 death but not severe COVID-19 (Figure 2 (W2)).

Results for the separate severe COVID-19 outcomes (death and hospitalisation), for the detailed (16-level) ethnicity analysis and for the analysis of the impact of household size are provided in the supplementary material.

There was no evidence of deviations from the proportional hazards assumption for wave 1 or wave 2 (Table S10).

### Other household-level variables and severe COVID-19 by ethnicity

There was strong evidence that the effect of deprivation on severe COVID-19 was different for White 67+ year olds compared to South Asian 67+ year olds in wave 2 (IMD most deprived vs least deprived - White: HR 1·65 95% CI 1·58-1·72, South Asian: HR 2·46 95% CI 2·00-3·03) (Table S10). This difference was specific to severe COVID-19 (Table S11).

### Absolute effects

Over 13% of South Asian people lived in multigenerational households in the most deprived settings (i.e. 67+ & 3 generations in the 5th deprivation quintile) compared to less than 1% of White people (Table 2). This meant that despite South Asian people making up only 3·2% of the total study population (compared to 94·7% White) (Table 1), there were a larger number of South Asian 67+ year olds (2841) than White 67+ year olds (2377) living in the highest risk household composition arrangement. South Asian people in this group experienced nearly triple the rate of severe COVID-19 (11802 per 100 000 person years) than White people in this group (4293 per 100 000 person-years) (Table 2), and the rate of severe COVID-19 for South Asian people in this group was higher than in those with two or more comorbidities (9905 per 100 000 person years) (Table 2).

**Table 2:** Rates of severe COVID by household composition, household size and deprivation status during Wave 2 for White and South Asian ethnicities (with figures for number of comorbidities included for comparison)

| *Wave 2 (1<sup>st</sup> September – 31<sup>st</sup> January 2021)* |  |  |  |  |
| --- | --- | --- | --- | --- |
|  | **N (%)** | **Events** | **Person years follow-up** | **Rate (per 100 000 person years)** |
| **White** |  |  |  |  |
| Index of Multiple Deprivation (IMD) |  |  |  |  |
| 1 (least deprived) | 658568 (25.6) | 5604 | 272071 | 2060 |
| 2 | 623364 (24.2) | 6018 | 257287 | 2339 |
| 3 | 550578 (21.4) | 6059 | 227038 | 2669 |
| 4 | 435765 (17.0) | 6173 | 179315 | 3443 |
| 5 (most deprived) | 302301 (11.8) | 5919 | 123922 | 4776 |
| Generational household composition |  |  |  |  |
| Multiple 67+ year olds | 1116533 (43.2) | 9893 | 461106 | 2145 |
| 67+ living alone | 880434 (34.1) | 14078 | 361731 | 3892 |
| 67+ & 1 other generation | 489303 (18.9) | 4932 | 201980 | 2442 |
| 67+ & 2 other generations | 84368 (3.3) | 847 | 34845 | 2431 |
| 67+ & 3 other generations | 12756 (0.5) | 173 | 5257 | 3291 |
| IMD and household composition |  |  |  |  |
| IMD=5, 67+ & 3 other generations | 2377 (0.8) | 42 | 978 | 4293 |
| Household size (number of occupants) |  |  |  |  |
| 1-2 | 2245408 (86.9) | 26269 | 925456 | 2838 |
| 3-5 | 314342 (12.2) | 3356 | 129712 | 2587 |
| 6+ | 23644 (0.9) | 298 | 9750 | 3056 |
| IMD and household size |  |  |  |  |
| IMD=5, HH size=6+ | 3471 (1.1) | 60 | 1428 | 4202 |
| Number of comorbidities (for comparison) |  |  |  |  |
| 2 or more | 684355 (26.5) | 17158 | 278833 | 6154 |
| **South Asian** |  |  |  |  |
| Index of Multiple Deprivation (IMD) |  |  |  |  |
| 1 (least deprived) | 9897 (11.1) | 108 | 4088 | 2642 |
| 2 | 12422 (13.9) | 214 | 5123 | 4177 |
| 3 | 19307 (21.6) | 383 | 7946 | 4820 |
| 4 | 25963 (29.1) | 653 | 10656 | 6128 |
| 5 (most deprived) | 21713 (24.3) | 725 | 8871 | 8173 |
| Generational household composition |  |  |  |  |
| Multiple 67+ year olds | 15969 (17.8) | 276 | 6572 | 4200 |
| 67+ living alone | 14745 (16.4) | 340 | 6064 | 5607 |
| 67+ & 1 other generation | 27605 (30.8) | 586 | 11355 | 5161 |
| 67+ & 2 other generations | 23136 (25.8) | 604 | 9494 | 6362 |
| 67+ & 3 other generations | 8198 (9.1) | 285 | 3345 | 8520 |
| IMD and household composition |  |  |  |  |
| IMD=5, 67+ & 3 other generations | 2841 (13.1) | 136 | 1152 | 11802 |
| Household size (number of occupants) |  |  |  |  |
| 1-2 | 38953 (43.4) | 799 | 16019 | 4988 |
| 3-5 | 31918 (35.6) | 672 | 13133 | 5117 |
| 6+ | 18782 (20.9) | 620 | 7678 | 8075 |
| IMD and Household size |  |  |  |  |
| IMD=5, HH size=6+ | 5594 (25.8) | 245 | 2271 | 10790 |
| Number of comorbidities (for comparison) |  |  |  |  |
| 2 or more | 35051 (39.1) | 1411 | 14246 | 9905 |

### Sensitivity analyses

All sensitivity analyses had minimal impact on results (Table S12).

## Discussion

### Principal findings

Multigenerational living was associated with increased risk of severe COVID-19 among older South Asian and White people in waves 1 and 2 of the SARS-CoV-2 pandemic in England, with evidence for a trend in increasing risk of severe COVID-19 with increasing number of generations in wave 2. For other ethnicities, results were consistent with harmful effects for wave 1 only. For South Asian older people, results were highly specific to COVID-19 and the effect of deprivation on severe COVID-19 outcomes was greater in South Asian than in White people. Very high rates of severe COVID-19 were observed for older people in multigenerational households in the most deprived settings. Despite there being over ten times the number of White people in England than South Asian people, nearly twice as many South Asian people live in multigenerational households in the most deprived settings compared to White people.

### Comparison with other studies

Our findings are consistent with two studies of older people (from the UK and from Sweden) that analysed COVID-19 mortality alone ^6,11^ and found increased risks associated with multigenerational living. Another UK study found that living with children was a risk factor for COVID-19 mortality in wave 2,^12^ and a study in the UK Biobank found an increased risk of severe COVID-19 in larger households during combined wave 1 and 2 (in younger people than studied here).^16^ A number of studies have reported ethnic disparities in COVID-19 outcomes in the UK.^17–21^ Of these, none studied age-based generational household composition by ethnic group and only two studies analysed effects over both of the first two waves of the pandemic.^17,21^

Our study complements and advances these previous studies by using both up-to-date household composition and covariate data from a very large population-representative sample of England to illustrate that as the number of generations in a household increased, the risk of severe COVID-19 for older people increased, and that effects were particularly pronounced for White and South Asian ethnicities during wave 2. Absolute rates of severe COVID-19 in wave 2 were very high for people living in multigenerational houses in the most deprived settings, and particularly for South Asian older people.

### Strengths and limitations

Our study is the first to use up-to-date household and covariate information to study household composition and severe COVID-19 in England by ethnicity, and was able to analyse effects over the first two waves where lockdown restrictions differed. The key strengths of this study are the scale, detail and completeness of the underlying health record data. We could therefore assess whether there was an increasing trend for COVID-19 harms in older people living with increasing numbers of generations by ethnicity, and assess the impact of other potential household-level explanatory variables (household size and deprivation).

Our study had a number of potential weaknesses. Firstly, 12% of our cohort who did not have a household identifier were excluded. Furthermore, for the main analysis 22% of the cohort were not included due to missing ethnicity data, although conclusions were identical when applying multiple imputation to account for missing ethnicity and the distribution of household composition classification by ethnicity was similar to Census data from 2011 (Table S13). There may have been some misclassification of household composition due to including some houses where not all occupants were registered at GP practices using TPP software, although our sensitivity analysis including only 100% TPP households had no impact on results. We were not able to account for people moving house during the pandemic, as household occupancy was determined only once (in February 2020). Finally, there were a number of potential explanatory factors that we could not include in this analysis, such as occupation^22^ and household crowding.

### Interpretation

Our results suggest that during wave 1 there were harmful effects of living with younger generations that were specific to COVID-19 for South Asian and other ethnicities, but no different to the effect on non-COVID-19 death for White older people. Possible explanations include the (non-specific) health benefits of multigenerational living,^10,23^ that strict lockdown intervention messaging was particularly effective at reaching White people,^24^ or that younger White people were more able to adhere to the messaging due to (e.g.) type of occupation. Older White people in households with more generations may have been less impacted specifically by COVID-19 as they had younger co-occupants who were able to provide support to the older occupant but were not coming into contact with other people outside of their own households. For the other ethnic groups, there may have been more contact between households (or in workplaces) during wave 1.^24^

For wave 2, increased multigenerational living had a large and specific effect on severe COVID-19 for South Asian older people. It is likely that as restrictions eased there were differences by ethnicity in key risk factors for transmission such as occupation type,^25^ inter-household mixing, religion, and experiences of structural racism.^17^ This could have led to increased exposure for South Asian people at work (or in education), and also for any South Asian older people in multigenerational houses. The fact that the effect of deprivation on severe COVID-19 was greater for South Asian older people than for White people provides further confirmation that differences in COVID-19 outcomes by ethnicity cannot be explained by deprivation alone.^26^

Finally, although the relative effects of multigenerational living were similar across White and South Asian groups during wave 2, the absolute effect differed substantially, with over one and a half times more people living in the highest risk household/deprivation category (n=5587 South Asian vs n=3415 White). As this group experienced over twice the rate of severe COVID-19 than their White counterparts and the rate was comparable to that in people with multiple established comorbidity risk factors, it is highly likely that these household-level characteristics are contributing to previously observed imbalances in severe COVID-19 outcomes by ethnicity that particularly impact South Asian people.^17,21^

## Conclusions

Multigenerational living was associated with severe COVID-19, particularly for South Asian and (to a lesser extent) White older people in both waves, with results consistent with harmful effects for other ethnicities in wave 1 only. The established COVID-19 risk factor of deprivation has a greater effect on serious COVID-19 outcomes for South Asian older people than for White older people. Older people in households with more than 3 other younger generations (or with six or more occupants) in the most deprived settings experienced particularly high rates of severe COVID-19. Household characteristics, ethnicity and socioeconomic deprivation are important considerations alongside individual risk factors when considering the roll-out of COVID-19 vaccination boosters and the targeting of interventions for COVID-19 and future pandemics.

## Data Availability

All data were linked stored and analysed securely within the OpenSAFELY-TPP platform: https://opensafely.org/. Data include pseudonymised data such as coded diagnoses medications and physiological parameters. No free text data are included. All code is shared openly for review and re-use under MIT open license [https://github.com/opensafely/hh-classification-research]. Detailed pseudonymised patient data is potentially re-identifiable and therefore not shared.

## Acknowledgements

We are very grateful for all the support received from the EMIS and TPP Technical Operations team throughout this work, and for generous assistance from the information governance and database teams at NHS England / NHSX.

## Data access and verification

Access to the underlying identifiable and potentially re-identifiable pseudonymised electronic health record data is tightly governed by various legislative and regulatory frameworks, and restricted by best practice. The data in OpenSAFELY is drawn from General Practice data across England where TPP is the Data Processor. TPP developers initiate an automated process to create pseudonymised records in the core OpenSAFELY database, which are copies of key structured data tables in the identifiable records. These are linked onto key external data resources that have also been pseudonymised via SHA-512 one-way hashing of NHS numbers using a shared salt. DataLab developers and PIs holding contracts with NHS England have access to the OpenSAFELY pseudonymised data tables as needed to develop the OpenSAFELY tools. These tools in turn enable researchers with OpenSAFELY Data Access Agreements to write and execute code for data management and data analysis without direct access to the underlying raw pseudonymised patient data, and to review the outputs of this code. All code for the full data management pipeline—from raw data to completed results for this analysis—and for the OpenSAFELY platform as a whole is available for review at https://github.com/opensafely/hh-classification-research.

The data management and analysis code for this paper was led by KW and contributed to by DG, RM, HPG, EN, AS, HF, KB, EW, HJC, CTR, AYSW.

## References

1. Li W, Zhang B, Lu J, Liu S, Chang Z, Peng C, et al. Characteristics of Household Transmission of COVID-19. Clin Infect Dis. 2020 Nov 5;71(8):1943–6.

2. Jing QL, Liu MJ, Zhang ZB, Fang LQ, Yuan J, Zhang AR, et al. Household secondary attack rate of COVID-19 and associated determinants in Guangzhou, China: a retrospective cohort study. Lancet Infect Dis. 2020 Oct 1;20(10):1141–50.

3. Li F, Li YY, Liu MJ, Fang LQ, Dean NE, Wong GWK, et al. Household transmission of SARS-CoV-2 and risk factors for susceptibility and infectivity in Wuhan: a retrospective observational study. Lancet Infect Dis. 2021 May 1;21(5):617–28.

4. Tsang TK, Lau LLH, Cauchemez S, Cowling BJ. Household Transmission of Influenza Virus. Trends Microbiol. 2016 Feb;24(2):123–33.

5. Endo A, Uchida M, Kucharski AJ, Funk S. Fine-scale family structure shapes influenza transmission risk in households: Insights from primary schools in Matsumoto city, 2014/15. PLOS Comput Biol. 2019 Dec 26;15(12):e1007589.

6. Brandén M, Aradhya S, Kolk M, Härkönen J, Drefahl S, Malmberg B, et al. Residential context and COVID-19 mortality among adults aged 70 years and older in Stockholm: a population-based, observational study using individual-level data. Lancet Healthy Longev. 2020 Nov 1;1(2):e80–8.

7. Rostila M, Cederström A, Wallace M, Brandén M, Malmberg B, Andersson G. Disparities in Coronavirus Disease 2019 Mortality by Country of Birth in Stockholm, Sweden: A Total-Population-Based Cohort Study. Am J Epidemiol. 2021 Aug 1;190(8):1510–8.

8. Stokes JE, Patterson SE. Intergenerational Relationships, Family Caregiving Policy, and COVID-19 in the United States. J Aging Soc Policy. 2020;32(4–5):416–24.

9. Haroon S, Chandan JS, Middleton J, Cheng KK. Covid-19: breaking the chain of household transmission. BMJ. 2020 Aug 14;370:m3181.

10. Arpino B, Bordone V, Pasqualini M. No clear association emerges between intergenerational relationships and COVID-19 fatality rates from macro-level analyses. Proc Natl Acad Sci U S A. 2020 Aug 11;117(32):19116–21.

11. Nafilyan V, Islam N, Ayoubkhani D, Gilles C, Katikireddi SV, Mathur R, et al. Ethnicity, household composition and COVID-19 mortality: a national linked data study. J R Soc Med. 2021 Apr;114(4):182–211.

12. Forbes H, Morton CE, Bacon S, McDonald HI, Minassian C, Brown JP, et al. Association between living with children and outcomes from covid-19: OpenSAFELY cohort study of 12 million adults in England. BMJ. 2021 Mar 18;372:n628.

13. Schultze A, Bates C, Cockburn J, MacKenna B, Nightingale E, Curtis HJ, et al. Identifying Care Home Residents in Electronic Health Records - An OpenSAFELY Short Data Report [Internet]. Wellcome Open Research; 2021 [cited 2021 Nov 22]. Available from: https://wellcomeopenresearch.org/articles/6-90

14. Williamson EJ, Walker AJ, Bhaskaran K, Bacon S, Bates C, Morton CE, et al. Factors associated with COVID-19-related death using OpenSAFELY. Nature. 2020 Aug;584(7821):430–6.

15. Grint DJ, Wing K, Williamson E, McDonald HI, Bhaskaran K, Evans D, et al. Case fatality risk of the SARS-CoV-2 variant of concern B.1.1.7 in England, 16 November to 5 February. Eurosurveillance. 2021 Mar 18;26(11):2100256.

16. Gillies CL, Rowlands AV, Razieh C, Nafilyan V, Chudasama Y, Islam N, et al. Association between household size and COVID-19: A UK Biobank observational study. J R Soc Med. 2022 Feb 4;01410768211073923.

17. Mathur R, Rentsch CT, Morton CE, Hulme WJ, Schultze A, MacKenna B, et al. Ethnic differences in SARS-CoV-2 infection and COVID-19-related hospitalisation, intensive care unit admission, and death in 17 million adults in England: an observational cohort study using the OpenSAFELY platform. The Lancet. 2021 May 8;397(10286):1711–24.

18. Ayoubkhani D, Nafilyan V, White C, Goldblatt P, Gaughan C, Blackwell L, et al. Ethnic-minority groups in England and Wales—factors associated with the size and timing of elevated COVID-19 mortality: a retrospective cohort study linking census and death records. Int J Epidemiol. 2020 Dec 8;dyaa208.

19. Niedzwiedz CL, O’Donnell CA, Jani BD, Demou E, Ho FK, Celis-Morales C, et al. Ethnic and socioeconomic differences in SARS-CoV-2 infection: prospective cohort study using UK Biobank. BMC Med. 2020 May 29;18(1):160.

20. Aldridge RW, Lewer D, Katikireddi SV, Mathur R, Pathak N, Burns R, et al. Black, Asian and Minority Ethnic groups in England are at increased risk of death from COVID-19: indirect standardisation of NHS mortality data. Wellcome Open Res. 2020 Jun 24;5:88.

21. Bhaskaran K, Bacon S, Evans SJ, Bates CJ, Rentsch CT, MacKenna B, et al. Factors associated with deaths due to COVID-19 versus other causes: population-based cohort analysis of UK primary care data and linked national death registrations within the OpenSAFELY platform. Lancet Reg Health - Eur. 2021 Jul 1;6:100109.

22. Hawkins D. Differential occupational risk for COVID-19 and other infection exposure according to race and ethnicity. Am J Ind Med. 2020 Jun 15;10.1002/ajim.23145.

23. Muennig P, Jiao B, Singer E. Living with parents or grandparents increases social capital and survival: 2014 General Social Survey-National Death Index. SSM - Popul Health. 2017 Nov 10;4:71–5.

24. Martin CA, Jenkins DR, Minhas JS, Gray LJ, Tang J, Williams C, et al. Socio-demographic heterogeneity in the prevalence of COVID-19 during lockdown is associated with ethnicity and household size: Results from an observational cohort study. EClinicalMedicine [Internet]. 2020 Aug 1 [cited 2021 Nov 18];25. Available from: https://www.thelancet.com/journals/eclinm/article/PIIS2589-5370(20)30210-8/fulltext

25. Coronavirus (COVID-19) related deaths by occupation, England and Wales - Office for National Statistics [Internet]. [cited 2021 Nov 18]. Available from: https://www.ons.gov.uk/peoplepopulationandcommunity/healthandsocialcare/causesofdeath/bulletins/coronaviruscovid19relateddeathsbyoccupationenglandandwales/latest

26. Razai MS, Kankam HKN, Majeed A, Esmail A, Williams DR. Mitigating ethnic disparities in covid-19 and beyond. BMJ. 2021 Jan 15;372m4921.

