## Supplementary Materials for "Association between household composition and severe COVID-19 outcomes in older people by ethnicity: an observational cohort study using the OpenSAFELY platform"

#### Supplementary material for Methods

##### OpenSAFELY - description of data source and information governance

**Data source**
All data were linked, stored and analysed securely within the OpenSAFELY-TPP platform: https://opensafely.org/. Data include pseudonymised data such as coded diagnoses, medications and physiological parameters. No free text data are included. All code is shared openly for review and re-use under MIT open license [https://github.com/opensafely/hh-classification-research]. Detailed pseudonymised patient data is potentially re-identifiable and therefore not shared.

Primary care records managed by the GP software provider, TPP were linked to NHS hospital records and ONS death data through OpenSAFELY-TPP.

**Information governance and ethical approval**
NHS England is the data controller for OpenSAFELY-TPP; TPP is the data processor; all study authors using OpenSAFELY have the approval of NHS England. This implementation of OpenSAFELY is hosted within the TPP environment which is accredited to the ISO 27001 information security standard and is NHS IG Toolkit compliant[1,2]

Patient data has been pseudonymised for analysis and linkage using industry standard cryptographic hashing techniques; all pseudonymised datasets transmitted for linkage onto OpenSAFELY are encrypted; access to the platform is via a virtual private network (VPN) connection, restricted to a small group of researchers; the researchers hold contracts with NHS England and only access the platform to initiate database queries and statistical models; all database activity is logged; only aggregate statistical outputs leave the platform environment following best practice for anonymisation of results such as statistical disclosure control for low cell counts.[3]

The OpenSAFELY research platform adheres to the obligations of the UK General Data Protection Regulation (GDPR) and the Data Protection Act 2018. In March 2020, the Secretary of State for Health and Social Care used powers under the UK Health Service (Control of Patient Information) Regulations 2002 (COPI) to require organisations to process confidential patient information for the purposes of protecting public health, providing healthcare services to the public and monitoring and managing the COVID-19 outbreak and incidents of exposure; this sets aside the requirement for patient consent.[4]

Taken together, these provide the legal bases to link patient datasets on the OpenSAFELY platform. GP practices, from which the primary care data are obtained, are required to share relevant health information to support the public health response to the pandemic, and have been informed of the OpenSAFELY analytics platform.

Health Research Authority (REC reference 20/LO/0651) and by the LSHTM Ethics Board (reference 21863).

**Algorithm to identify households in TPP, using addresses registered in patient record**
1) Get all address details for fully registered patients - Find all permanent patient addresses that were active on 01 Feb 2020 or recorded since - Combine the house name/number, remove punctuation (commas, full stops, apostrophes), double spaces and leading/trailing whitespace from house name/number and road - Replace street, lane etc with the abbreviation (street -> St, lane -> Ln, place -> Pl, avenue -> Ave, road -> Rd, close -> Cl, drive -> Dr) - Find registrations for patients that were active on or since 01 Feb 2020 (making sure the patient was alive on 01 Feb 2020) - Filter the patient addresses based on those registrations
2) Build an Address table using the distinct address fields - For each distinct (house name/number + road + post code), assign a unique Address_ID
3) Set the Address_ID on each individual patient address - Join to Address table from 2) on house name/number + road + post code
4) End addresses that started before the property was sold - Import land registry property sales data (www.gov.uk/government/statistical-data-sets/price-paid-data- downloads) - Apply the same processing as 1) to the address fields - Set address IDs on the land registry house sales data using the same Address table from 2)

joining on house name/number + road + post code - End all patient addresses at properties where the patient address start date was before the property was sold (under the assumption that in the majority of cases that means the occupants moved out) - If a patient address has an unset start date (imported data), use the registration start date for this step

5) Get the latest "active" patient address per patient - Filter to those which were active on 01 Feb 2020 or recorded since (need to do this again now we've ended some based on property sales) - Filter to the latest per patient

6) Create the HouseholdMember table - Insert a row for each patient address from 5), using the Address_ID as the Household_ID (one household per address)

7) Create the Household table - Select the distinct Address_IDs as the Household_ID into the Household table (one household per address) - Set the size based on the number of members - Use the PotentialCareHomeAddress table to set the CareHome flag - Check the address fields to identify NFA / Unknown addresses (e.g. postcode "ZZ99 ...", house name "NFA", road "Unknown", ...)

**Analysis of the impact of household size (number of occupants)**

In our main analysis of household composition we did not adjust for household size (number of occupants) as conceptually household size is a mediator of household composition. We also noted that not all categories of the household composition variable had a corresponding household size category (Supplementary Table S5), meaning that adjusting for household size would artificially reduce estimates downwards.

**Household TPP coverage**

In England, not all residents of a household are necessarily registered with the same general practice, which means it is possible that not all residents of a household would appear in the same software system. This means that the number of residents attributed to a household in the TPP register is not necessarily equal to the true total number of people in the household.

While all households in our study will have included at least one 67+ year old registered at a practice using TPP software, it is possible that some of the other people in some of the households may be registered with other practices that use software other than TPP. For these households, the calculated TPP household size (i.e. the count of records in TPP under the same household ID) will be different from the household size defined in the Master Patient Index (MPI) for the address covered by the TPP household ID. Those people who are registered to non-TPP practices will not have been counted when we created our household composition exposure variable, and this measurement error has the potential to bias results. A "TPP coverage" flag is provided for each household, which compares the TPP household size with the number of records in the Master Patient Index for the same address and is used to indicate the % of occupants of that household who are registered with general practices that use TPP.

For our main analysis, we included all households, irrespective of TPP coverage. In order to assess the impact of this design choice on our results, we performed a sensitivity analysis where we only included households with 100% TPP coverage.

#### Table S1: Deviations from the original study protocol

| Deviation | Rationale |
|---|---|
| Our main exposure was categorised into 5 categories rather than 3 as detailed in the protocol | This change was made to facilitate analysis for a test for trend of increasing/decreasing severe COVID with increasing number of generations. |
| Our primary outcome in the analysis was a combined outcome of COVID death or hospitalisation. In the protocol we specified these as separate outcomes but not as a combined outcome. | This additional outcome definition was included in order to increase power to detect effects in all ethnic groups analysed. |
| We included a step to compare the distribution of household sizes by ethnicity with ONS figures from the 2011 census. | This was to ensure that the TPP method for assigning people to households was correct. |
| We added a number of ways of identifying people in care homes to the final analysis (vs only a single method in the protocol). | This change was made based on best practice for identifying care homes (Schultze et al 2021) that was published during preparation of our own analysis. |
| In the analysis we looked for evidence of interaction between ethnicity and any other variable, and if there were interactions found between ethnicity and any household-level variable (i.e. deprivation or housing density) presented strata for these, whereas in the protocol we only said we would stratify by deprivation. | This was to ensure we did not miss any potentially important interactions with ethnicity. |

#### Table S2: Table showing high-level ethnicity with component ethnicity categories

| High level ethnicity categories | Component categories |
|---|---|
| White | White British, White Irish, other White |
| South Asian | Indian, Pakistani, Bangladeshi, other South Asian |
| Black | African, Caribbean, other Black |
| Mixed | White and Asian, White and African, White and Caribbean, other mixed |
| Other | Chinese, all others |

**Table S3: Table of covariate definitions**

| Covariate | Description |
|---|---|
| Sex | Male or female |
| Age | Restricted cubic splines were used for the main analysis of each ethnicity cohort. When grouped (for descriptive reporting and when including interaction with age in the combined ethnicity cohort) age groups were: 67-69, 70-74, 75-79, 80-84, 85+. |
| Ethnicity | Categorised as a five-level variable based on primary care self reporting of: White, Mixed, South Asian, Black or Other |
| BMI | Body mass index (BMI) was ascertained within the 10 years prior to 1 Feb 2020 and recorded when the patient was over 16 years old. Grouped using categories derived from the World Health Organisation classification of BMI: underweight, normal, overweight, obese I 30-34.9; obese II 35-39.9; obese III 40+. Those with missing BMI were assigned to the normal weight category. |
| Smoking status | Grouped into current, former and never smokers. Missing smoking data were assigned to the never smokers category. |
| Index of Multiple Deprivation | Definition of deprivation using quintiles of the Index of Multiple Deprivation, an area-level composite measure of seven domains: income, employment, education, skills and training, health and disability, crime, and barriers to housing services and living environment.[6] |
| Region of England | Nine categories: East, East Midlands, London, North East, North West, South East, South West, West Midlands, Yorkshire & The Humber |
| Geographic area | ONS UTLA (Upper Tier Local Authority) - used as a stratification variable |
| Housing density | ONS Rural/Urban classification, divided into five categories: urban major conurbation, urban minor conurbation, urban city and town, rural town, rural village. |
| Total number of people in the household | Categorised as 1-2, 3-5, 6 or more. |
| Comorbidities | Presence of either 0, 1 or 2 or more comorbidities (see Table S3 below for further details) |

**Table S4: Definitions of comorbidities**

Based on previous work on association of specific comorbidities with severe COVID-19,[5] we included a comorbidity variable with categories capturing whether each 67+ year old included in the study had either 0, 1 or 2+ comorbidities recorded prior to their start of follow-up. The list of conditions and codelists that were used to define the presence of comorbidity are provided below.

| Condition | Codelist defining presence of comorbidity |
|---|---|
| Aplastic anaemia | https://codelists.opensafely.org/codelist/opensafely/aplastic-anaemia/ |
| Asplenia | https://codelists.opensafely.org/codelist/opensafely/asplenia/ |
| Asthma | https://codelists.opensafely.org/codelist/opensafely/asthma-diagnosis/<br>https://codelists.opensafely.org/codelist/opensafely/asthma-inhaler-salbutamol-medication/2020-04-15/<br>https://codelists.opensafely.org/codelist/opensafely/asthma-inhaler-steroid-medication/2020-04-15/<br>https://codelists.opensafely.org/codelist/opensafely/asthma-oral-prednisolone-medication/2020-04-27/ |
| Bone marrow transplant | https://codelists.opensafely.org/codelist/opensafely/bone-marrow-transplant/2020-04-15/ |
| Cancer | https://codelists.opensafely.org/codelist/opensafely/cancer-excluding-lung-and-haematological/2020-04-15/<br>https://codelists.opensafely.org/codelist/opensafely/chemotherapy-or-radiotherapy-updated/2020-04-15/<br>https://codelists.opensafely.org/codelist/opensafely/haematological-cancer/2020-04-15/<br>https://codelists.opensafely.org/codelist/opensafely/lung-cancer/2020-04-15/ |
| Chronic cardiac disease | https://codelists.opensafely.org/codelist/opensafely/chronic-cardiac-disease/2020-04-08/ |
| Chronic respiratory disease | https://codelists.opensafely.org/codelist/opensafely/chronic-respiratory-disease/2020-04-10/ |
| Chronic liver disease | https://codelists.opensafely.org/codelist/opensafely/chronic-liver-disease/2020-06-02/ |
| Dementia | https://codelists.opensafely.org/codelist/opensafely/dementia/2020-04-22/ |
| Diabetes | https://codelists.opensafely.org/codelist/opensafely/diabetes/2020-04-15/ |
| Chronic kidney disease | https://codelists.opensafely.org/codelist/opensafely/chronic-kidney-disease/2020-04-14/ |
| GI bleed | https://codelists.opensafely.org/codelist/opensafely/gi-bleed-or-ulcer/2020-04-08/ |
| HIV | https://codelists.opensafely.org/codelist/opensafely/hiv/2020-07-13/ |
| Permanent immunosuppression | https://codelists.opensafely.org/codelist/opensafely/permanent-immunosuppression/2020-06-02/ |
| Temporary immunosuppression | https://codelists.opensafely.org/codelist/opensafely/temporary-immunosuppression/2020-04-24/ |
| Hypertension | https://codelists.opensafely.org/codelist/opensafely/hypertension/2020-04-28/ |
| Stroke | https://codelists.opensafely.org/codelist/opensafely/stroke-updated/2020-06-02/ |
| Inflammatory bowel disease | https://codelists.opensafely.org/codelist/opensafely/inflammatory-bowel-disease/2020-04-07/ |
| Neurological conditions | https://codelists.opensafely.org/codelist/opensafely/other-neurological-conditions/2020-06-02/ |
| Psoriasis | https://codelists.opensafely.org/codelist/opensafely/ra-sle-psoriasis/2020-04-14/ |
| Sickle cell disease | https://codelists.opensafely.org/codelist/opensafely/sickle-cell-disease/2020-04-14/ |
| Smoking | https://codelists.opensafely.org/codelist/opensafely/smoking-clear/2020-04-29/<br>https://codelists.opensafely.org/codelist/opensafely/smoking-unclear/2020-04-29/ |
| Organ transplant | https://codelists.opensafely.org/codelist/opensafely/solid-organ-transplantation/2020-04-10/ |

**Table S5: Table showing that not all categories of household composition have a corresponding household size category**

| Household composition category | Possible household size | | | | | |
|---|---|---|---|---|---|---|
| | 1 | 2 | 3 | 4 | 5 | 6+ |
| Multiple 67+ year olds (max 4*) | ✗ | ✓ | ✓ | ✓ | ✗ | ✗ |
| 67+ living alone | ✓ | ✗ | ✗ | ✗ | ✗ | ✗ |
| 67+ & 1 other generation | ✗ | ✓ | ✓ | ✓ | ✓ | ✓ |
| 67+ & 2 other generations | ✗ | ✗ | ✓ | ✓ | ✓ | ✓ |
| 67+ & 3 other generations | ✗ | ✗ | ✗ | ✓ | ✓ | ✓ |

*largest allowable size of house with only 67+ year olds in in this study

**Table S6: Categories for variable combining household size and household composition**

| Combined hh composition - hh size variable |
|---|
| Two 67+ year olds (hhsize=2) |
| >2 67+ year olds (hhsize=3-4) |
| 67+ & 1 gen (hhsize=2) |
| 67+ & 1 gen (hhsize=3-4) |
| 67+ & 1 gen (hhsize=5+) |
| 67+ & 2 gen (hhsize=3-4) |
| 67+ & 2 gen (hhsize=5+) |
| 67+ & 3 gen (hhsize=3-4) |
| 67+ & 3 gen (hhsize=5+) |

**Software and Reproducibility**

This analysis was delivered through the OpenSAFELY platform: codelists and code for data management and data analysis were specified using the OpenSAFELY tools; then transmitted securely to the OpenSAFELY-TPP platform within TPP's secure environment, where they were executed against local patient data; summary results were then reviewed for disclosiveness, released, and formatted for the final outputs. All code for the OpenSAFELY platform for data management, analysis and secure code execution is shared for review and re-use under open licenses at GitHub.com/OpenSAFELY. Data management and analysis was performed using Python 3.8 and Stata 16.1. Code for data management and analysis as well as codelists archived online https://github.com/opensafely/hh-classification-research.

#### Supplementary material for Results

**Figure S1: Flow diagram of cohort with numbers excluded at different stages for wave 1 and wave 2**

**Wave 1**

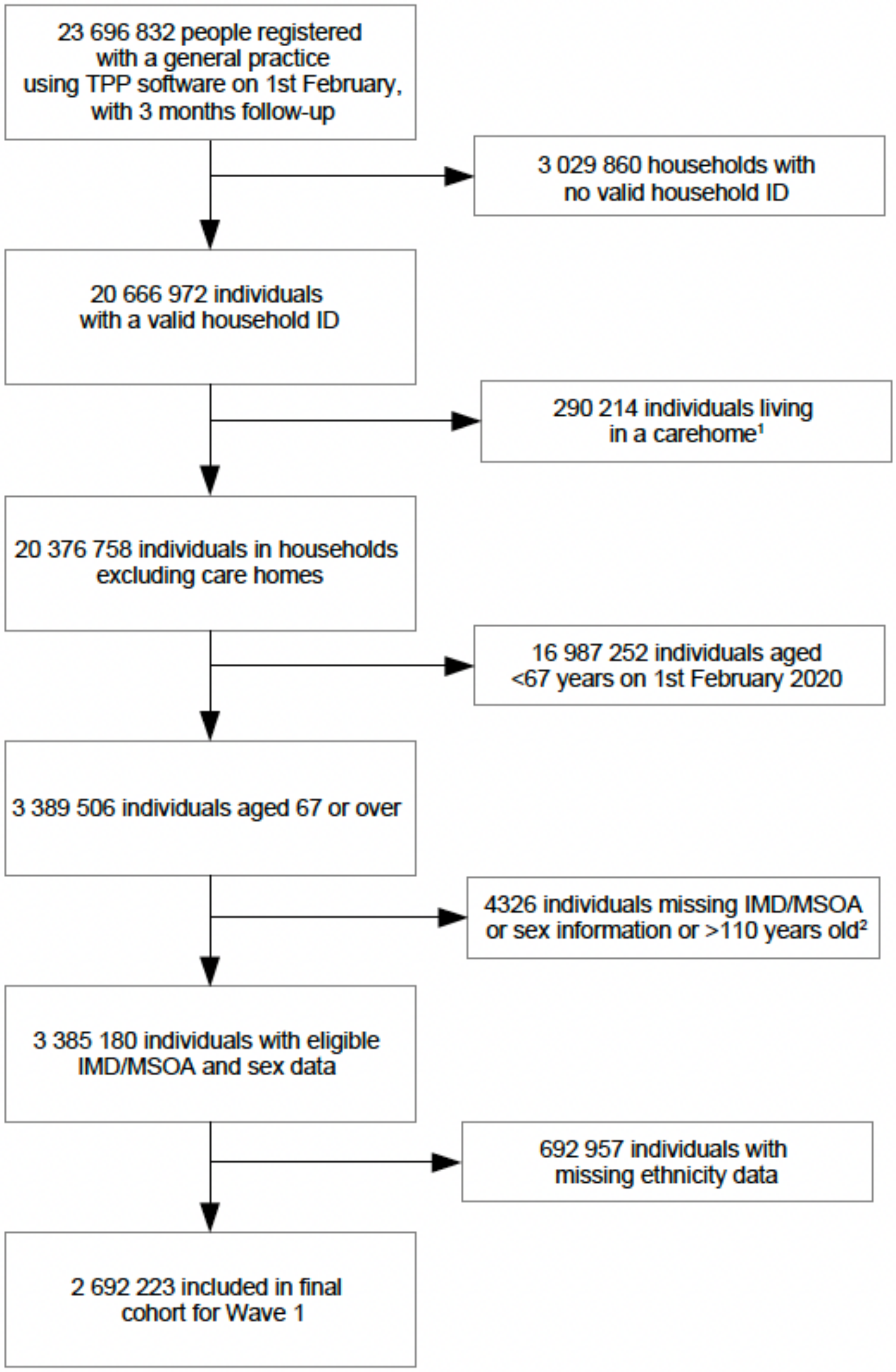


**Note 1:** 97 143 flagged as living in a care home, 13 999 living in a house that had an individual flagged as living in a care home, 176 701 living in a private home >12 in size, 2 371 living in a private home >4 in size with all occupants >67.
**Note 2:** 4 312 missing IMD/MSOA or sex information, 14 aged>110 years

**Wave 2**

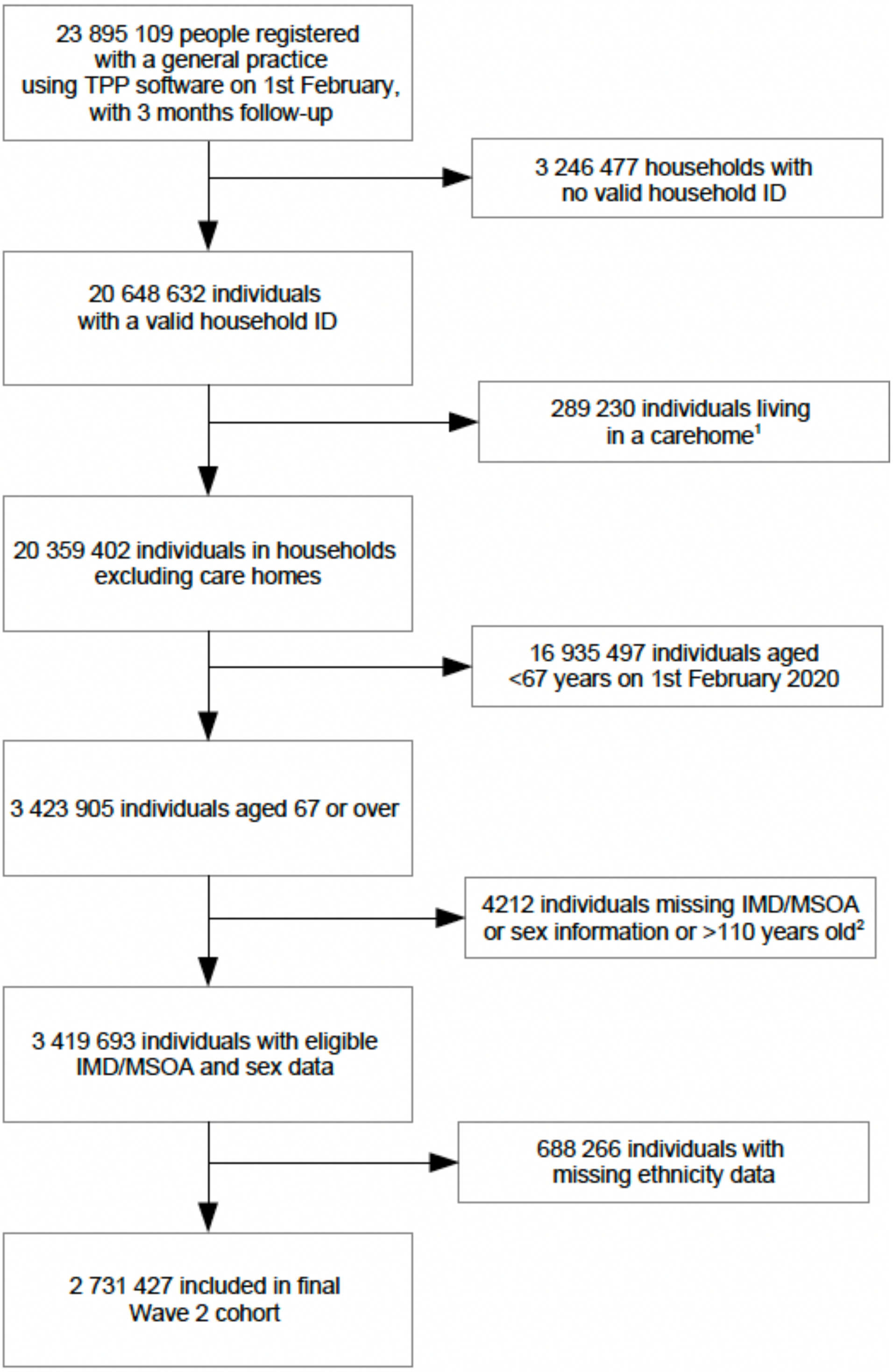


**Note 1:** 93 548 flagged as living in a care home, 27 420 living in a house that had an individual flagged as living in a care home, 166 723 living in a private home >12 in size, 1 539 living in a private home >4 in size with all occupants >67.
**Note 2:** 4 198 missing IMD/MSOA or sex information, 14 aged>110 years

**Table S7: Association between household composition (number of generations in a household) and severe COVID (hospitalisation or death as a combined outcome) or non-COVID death in 67+ year olds, for separate ethnicity cohorts during the first and second waves of the pandemic in the UK, with serial adjustment**

*A. Death or hospitalisation from COVID-19 (combined outcome)*

| | Wave 1 (1st February – 31st August 2020) | | | | | | Wave 2 (1st September – 31st January 2021) | | | | | |
|---|---|---|---|---|---|---|---|---|---|---|---|---|
| | | | | HR (95% CI) | | | | | | HR (95% CI) | | |
| HH composition[1] | N (%) | P-years | Rate[2] | Crude[3] | Age-adjusted[3] | Plus sex, deprivation, comorbidities clinical factors[4] | N (%) | P-years | Rate[2] | Crude[3] | Age-adjusted[3] | Plus sex, deprivation, comorbidities clinical factors[5] |
| White | | | | | | | | | | | | |
| Multiple 67+ | 1083552 (42·5) | 622902 | 736 | 1 | 1 | 1 | 1116533 (43·2) | 461106 | 2145 | 1 | 1 | 1 |
| 67+ alone | 887343 (34·8) | 506608 | 1317 | 1·71 (1·64-1·78) | 1·33 (1·28-1·39) | 1·35 (1·30-1·41) | 880434 (34·1) | 361731 | 3892 | 1·76 (1·71-1·81) | 1·39 (1·35-1·43) | 1·37 (1·33-1·41) |
| 67+ & 1 gen | 482696 (18·9) | 277366 | 808 | 1·04 (0·98-1·09) | 1·21 (1·15-1·27) | 1·14 (1·08-1·20) | 489303 (18·9) | 201980 | 2442 | 1·10 (1·06-1·14) | 1·29 (1·24-1·34) | 1·22 (1·17-1·26) |
| 67+ & 2 gens | 82362 (3·2) | 47317 | 845 | 1·06 (0·95-1·17) | 1·28 (1·15-1·42) | 1·22 (1·10-1·35) | 84368 (3·3) | 34845 | 2431 | 1·09 (1·01-1·17) | 1·33 (1·24-1·43) | 1·26 (1·17-1·36) |
| 67+ & 3 gens | 12309 (0·5) | 7057 | 907 | 1·10 (0·86-1·40) | 1·28 (1·00-1·64) | 1·21 (0·94-1·55) | 12756 (0·5) | 5257 | 3291 | 1·44 (1·24-1·68) | 1·70 (1·46-1·97) | 1·61 (1·38-1·87) |
| South Asian | | | | | | | | | | | | |
| Multiple 67+ | 15264 (17·5) | 8741 | 1579 | 1 | 1 | 1 | 15969 (17·8) | 6572 | 4200 | 1 | 1 | 1 |
| 67+ alone | 14674 (16·8) | 8366 | 2510 | 1·53 (1·22-1·91) | 1·42 (1·14-1·77) | 1·47 (1·18-1·84) | 14745 (16·4) | 6064 | 5607 | 1·31 (1·11-1·55) | 1·23 (1·04-1·45) | 1·22 (1·03-1·44) |
| 67+ & 1 gen | 26557 (30·5) | 15195 | 1889 | 1·14 (0·92-1·41) | 1·20 (0·97-1·48) | 1·17 (0·95-1·45) | 27605 (30·8) | 11355 | 5161 | 1·19 (1·03-1·39) | 1·27 (1·09-1·47) | 1·22 (1·05-1·42) |
| 67+ & 2 gens | 22650 (26·0) | 12946 | 2186 | 1·33 (1·07-1·64) | 1·36 (1·09-1·68) | 1·34 (1·08-1·66) | 23136 (25·8) | 9494 | 6362 | 1·46 (1·26-1·70) | 1·51 (1·30-1·75) | 1·45 (1·25-1·69) |
| 67+ & 3 gens | 7987 (9·2) | 4560 | 2478 | 1·49 (1·15-1·92) | 1·45 (1·12-1·87) | 1·41 (1·09-1·83) | 8198 (9·1) | 3345 | 8520 | 1·94 (1·63-2·31) | 1·91 (1·60-2·27) | 1·76 (1·48-2·10) |
| Black | | | | | | | | | | | | |
| Multiple 67+ | 4019 (15·2) | 2295 | 2136 | 1 | 1 | 1 | 4132 (15·3) | 1701 | 5057 | 1 | 1 | 1 |
| 67+ alone | 9399 (35·5) | 5346 | 2918 | 1·25 (0·90-1·74) | 1·22 (0·88-1·70) | 1·24 (0·89-1·72) | 9462 (35·1) | 3889 | 5760 | 1·09 (0·84-1·41) | 1·07 (0·82-1·38) | 1·07 (0·82-1·39) |
| 67+ & 1 gen | 7888 (29·8) | 4506 | 2130 | 0·91 (0·64-1·30) | 0·95 (0·67-1·36) | 1·01 (0·71-1·44) | 8050 (29·9) | 3315 | 4194 | 0·80 (0·60-1·05) | 0·84 (0·63-1·11) | 0·88 (0·66-1·17) |
| 67+ & 2 gens | 4017 (15·2) | 2305 | 2213 | 0·97 (0·65-1·46) | 1·09 (0·73-1·62) | 1·27 (0·85-1·90) | 4119 (15·3) | 1700 | 3354 | 0·64 (0·45-0·90) | 0·72 (0·51-1·01) | 0·83 (0·59-1·17) |
| 67+ & 3 gens | 1155 (4·4) | 661 | 1966 | 0·85 (0·46-1·58) | 0·94 (0·51-1·74) | 1·16 (0·62-2·14) | 1185 (4·4) | 490 | 4281 | 0·82 (0·50-1·32) | 0·90 (0·56-1·47) | 1·11 (0·69-1·81) |
| Mixed | | | | | | | | | | | | |
| Multiple 67+ | 2027 (21·6) | 1162 | 1290 | 1 | 1 | 1 | 2117 (21·8) | 871 | 3903 | 1 | 1 | 1 |
| 67+ alone | 3129 (33·3) | 1784 | 1962 | 1·36 (0·71-2·63) | 1·35 (0·70-2·61) | 1·29 (0·67-2·49) | 3188 (32·9) | 1310 | 4198 | 1·02 (0·66-1·57) | 0·97 (0·62-1·50) | 0·90 (0·57-1·42) |
| 67+ & 1 gen | 2711 (28·9) | 1554 | 1480 | 1·01 (0·50-2·03) | 1·16 (0·58-2·32) | 1·16 (0·58-2·32) | 2832 (29·2) | 1169 | 2824 | 0·68 (0·42-1·10) | 0·70 (0·44-1·14) | 0·70 (0·43-1·15) |
| 67+ & 2 gens | 1209 (12·9) | 696 | 1437 | 0·96 (0·41-2·23) | 1·12 (0·48-2·60) | 1·24 (0·53-2·87) | 1234 (12·7) | 509 | 4318 | 1·01 (0·59-1·74) | 1·09 (0·63-1·86) | 1·12 (0·65-1·92) |
| 67+ & 3 gens | 318 (3·4) | 184 | 0 | 0·00 (0·00-0·00) | 0·00 (0·00-0·00) | 0·00 (0·00-0·00) | 325 (3·4) | 134 | 6736 | 1·53 (0·73-3·21) | 1·58 (0·76-3·30) | 1·62 (0·77-3·41) |
| Other | | | | | | | | | | | | |
| Multiple 67+ | 4581 (21·9) | 2635 | 721 | 1 | 1 | 1 | 4807 (22·1) | 1984 | 3478 | 1 | 1 | 1 |
| 67+ alone | 6125 (29·2) | 3505 | 1541 | 1·89 (1·10-3·25) | 1·72 (1·00-2·97) | 1·72 (0·99-2·97) | 6210 (28·6) | 2559 | 4299 | 1·18 (0·86-1·62) | 1·12 (0·82-1·54) | 1·10 (0·80-1·51) |
| 67+ & 1 gen | 6185 (29·5) | 3558 | 1237 | 1·54 (0·88-2·69) | 1·65 (0·94-2·89) | 1·66 (0·95-2·91) | 6486 (29·8) | 2682 | 2908 | 0·79 (0·56-1·11) | 0·86 (0·61-1·21) | 0·84 (0·60-1·19) |
| 67+ & 2 gens | 3295 (15·7) | REDACT | REDACT | REDACT | 1·75 (0·94-3·26) | 1·88 (1·01-3·50) | 3421 (15·7) | 1417 | 3034 | 0·82 (0·56-1·21) | 0·90 (0·61-1·33) | 0·96 (0·65-1·42) |
| 67+ & 3 gens | 771 (3·7) | REDACT | REDACT | REDACT | 0·87 (0·26-2·99) | 0·89 (0·26-3·07) | 812 (3·7) | 335 | 3876 | 1·03 (0·54-1·94) | 1·09 (0·58-2·07) | 1·12 (0·59-2·13) |

Note 1: Household composition in terms of number of distinct generations that each 67+ year old in the study cohort is living with (considering the following distinct generations: 0-17 year olds, 18-29 year olds, 30-66 year olds, 67+ year olds) i.e. 67+ & 1=67+ year old's houshold includes one other generation, 67+ & 2=67+ year old's houshold includes two other generations. Note 2: Rate per 100 000 person years. Note 3: Stratified by location (UTLA). Note 4: Stratified by location (UTLA) and adjusted for sex, number of comorbidities, categories of housing density (rural or urban setting), smoking status, socio-economic status and includng an interaction between ethnicity and age (as well as the interaction between household composition and ethnicity presented here). Note 5: Stratified on UTLA and adjusted for: sex, smoking, housing density and number of comorbidities and including interactions between ethnicity and: IMD, age and obesity (as well as the interaction with household composition presented here).

*B. Non-COVID-19 death*

| | Wave 1 (1st February – 31st August 2020) | | | | | | Wave 2 (1st September – 31st January 2021) | | | | | |
|---|---|---|---|---|---|---|---|---|---|---|---|---|
| | | | | HR (95% CI) | | | | | | HR (95% CI) | | |
| **HH composition[1]** | **N (%)** | **P-years** | **Rate[2]** | **Crude[3]** | **Age-adjusted[3]** | **Plus sex, deprivation, comorbidities clinical factors[4]** | **N (%)** | **P-years** | **Rate[2]** | **Crude[3]** | **Age-adjusted[3]** | **Plus sex, deprivation, comorbidities clinical factors[5]** |
| White | | | | | | | | | | | | |
| Multiple 67+ | 1083552 (42·5) | 623800 | 2390 | 1 | 1 | 1 | 1116533 (43·2) | 462132 | 2269 | 1 | 1 | 1 |
| 67+ alone | 887343 (34·8) | 507924 | 4036 | 1·67 (1·63-1·70) | 1·21 (1·19-1·24) | 1·19 (1·17-1·22) | 880434 (34·1) | 363217 | 3655 | 1·60 (1·55-1·64) | 1·17 (1·14-1·20) | 1·15 (1·12-1·18) |
| 67+ & 1 gen | 482696 (18·9) | 277844 | 2424 | 1·00 (0·97-1·03) | 1·20 (1·16-1·23) | 1·13 (1·10-1·16) | 489303 (18·9) | 202497 | 2294 | 1·00 (0·97-1·04) | 1·21 (1·17-1·26) | 1·16 (1·12-1·20) |
| 67+ & 2 gens | 82362 (3·2) | 47403 | 2456 | 1·02 (0·96-1·08) | 1·26 (1·19-1·34) | 1·18 (1·11-1·25) | 84368 (3·3) | 34931 | 2242 | 0·98 (0·91-1·06) | 1·24 (1·16-1·34) | 1·18 (1·10-1·27) |
| 67+ & 3 gens | 12309 (0·5) | 7072 | 2814 | 1·16 (1·00-1·33) | 1·37 (1·19-1·57) | 1·25 (1·09-1·44) | 12756 (0·5) | 5273 | 2731 | 1·19 (1·01-1·40) | 1·43 (1·22-1·69) | 1·34 (1·14-1·58) |
| South Asian | | | | | | | | | | | | |
| Multiple 67+ | 15264 (17·5) | 8766 | 2761 | 1 | 1 | 1 | 15969 (17·8) | 6601 | 2363 | 1 | 1 | 1 |
| 67+ alone | 14674 (16·8) | 8404 | 2927 | 1·07 (0·90-1·28) | 0·91 (0·76-1·09) | 0·97 (0·81-1·16) | 14745 (16·4) | 6096 | 2657 | 1·14 (0·92-1·43) | 0·97 (0·78-1·21) | 1·04 (0·83-1·30) |
| 67+ & 1 gen | 26557 (30·5) | 15247 | 2538 | 0·91 (0·78-1·07) | 0·99 (0·84-1·16) | 1·00 (0·85-1·18) | 27605 (30·8) | 11408 | 2410 | 1·02 (0·83-1·24) | 1·11 (0·91-1·36) | 1·13 (0·92-1·38) |
| 67+ & 2 gens | 22650 (26·0) | 12994 | 2609 | 0·93 (0·79-1·10) | 0·95 (0·80-1·12) | 0·98 (0·83-1·15) | 23136 (25·8) | 9554 | 2334 | 0·97 (0·79-1·19) | 1·00 (0·81-1·22) | 1·02 (0·83-1·26) |
| 67+ & 3 gens | 7987 (9·2) | 4578 | 3145 | 1·11 (0·90-1·36) | 1·02 (0·83-1·25) | 1·03 (0·84-1·27) | 8198 (9·1) | 3374 | 2875 | 1·18 (0·91-1·52) | 1·09 (0·84-1·40) | 1·12 (0·87-1·46) |
| Black | | | | | | | | | | | | |
| Multiple 67+ | 4019 (15·2) | 2304 | 2995 | 1 | 1 | 1 | 4132 (15·3) | 1707 | 2637 | 1 | 1 | 1 |
| 67+ alone | 9399 (35·5) | 5380 | 3476 | 1·16 (0·88-1·52) | 1·09 (0·83-1·44) | 1·09 (0·83-1·44) | 9462 (35·1) | 3908 | 2943 | 1·11 (0·79-1·57) | 1·04 (0·74-1·47) | 1·05 (0·74-1·49) |
| 67+ & 1 gen | 7888 (29·8) | 4527 | 2849 | 0·94 (0·70-1·26) | 1·01 (0·76-1·35) | 1·07 (0·80-1·43) | 8050 (29·9) | 3327 | 2765 | 1·05 (0·73-1·50) | 1·11 (0·78-1·59) | 1·17 (0·82-1·67) |
| 67+ & 2 gens | 4017 (15·2) | 2317 | 1338 | 0·45 (0·29-0·68) | 0·54 (0·35-0·82) | 0·66 (0·43-1·00) | 4119 (15·3) | 1705 | 2288 | 0·88 (0·57-1·34) | 1·05 (0·68-1·61) | 1·24 (0·80-1·91) |
| 67+ & 3 gens | 1155 (4·4) | 665 | 2556 | 0·85 (0·50-1·44) | 1·00 (0·59-1·70) | 1·29 (0·76-2·20) | 1185 (4·4) | 492 | 1219 | 0·46 (0·20-1·09) | 0·55 (0·23-1·29) | 0·57 (0·23-1·43) |
| Mixed | | | | | | | | | | | | |
| Multiple 67+ | 2027 (21·6) | 1164 | 2921 | 1 | 1 | 1 | 2117 (21·8) | 874 | 2518 | 1 | 1 | 1 |
| 67+ alone | 3129 (33·3) | 1793 | 3124 | 1·06 (0·69-1·62) | 0·98 (0·64-1·51) | 0·96 (0·62-1·46) | 3188 (32·9) | 1316 | 3040 | 1·21 (0·72-2·03) | 1·08 (0·64-1·83) | 1·04 (0·62-1·73) |
| 67+ & 1 gen | 2711 (28·9) | 1558 | 2118 | 0·71 (0·44-1·15) | 0·81 (0·50-1·30) | 0·82 (0·51-1·32) | 2832 (29·2) | 1172 | 2390 | 0·94 (0·54-1·64) | 1·03 (0·59-1·79) | 1·04 (0·60-1·80) |
| 67+ & 2 gens | 1209 (12·9) | REDACT | REDACT | REDACT | 0·39 (0·17-0·88) | 0·43 (0·19-0·97) | 1234 (12·7) | REDACT | REDACT | REDACT | 0·88 (0·42-1·83) | 0·86 (0·40-1·83) |
| 67+ & 3 gens | 318 (3·4) | REDACT | REDACT | REDACT | 0·41 (0·10-1·72) | 0·45 (0·11-1·89) | 325 (3·4) | REDACT | REDACT | REDACT | 0·61 (0·14-2·59) | 0·67 (0·16-2·84) |
| Other | | | | | | | | | | | | |
| Multiple 67+ | 4581 (21·9) | 2639 | 2274 | 1 | 1 | 1 | 4807 (22·1) | 1990 | 2262 | 1 | 1 | 1 |
| 67+ alone | 6125 (29·2) | 3516 | 2816 | 1·24 (0·90-1·71) | 1·05 (0·76-1·45) | 1·03 (0·74-1·42) | 6210 (28·6) | 2569 | 2180 | 0·98 (0·66-1·46) | 0·85 (0·56-1·28) | 0·85 (0·56-1·28) |
| 67+ & 1 gen | 6185 (29·5) | 3568 | 1597 | 0·69 (0·48-1·00) | 0·75 (0·52-1·08) | 0·77 (0·53-1·11) | 6486 (29·8) | 2689 | 1636 | 0·72 (0·47-1·09) | 0·83 (0·54-1·26) | 0·84 (0·55-1·28) |
| 67+ & 2 gens | 3295 (15·7) | 1905 | 420 | 0·18 (0·09-0·38) | 0·21 (0·10-0·43) | 0·23 (0·11-0·47) | 3421 (15·7) | REDACT | REDACT | REDACT | 0·47 (0·26-0·88) | 0·52 (0·28-0·96) |
| 67+ & 3 gens | 771 (3·7) | 445 | 2471 | 1·05 (0·55-1·99) | 1·15 (0·61-2·19) | 1·26 (0·66-2·37) | 812 (3·7) | REDACT | REDACT | REDACT | 0·70 (0·28-1·77) | 0·79 (0·31-2·00) |

Note 1: Household composition in terms of number of distinct generations that each 67+ year old in the study cohort is living with (considering the following distinct generations: 0-17 year olds, 18-29 year olds, 30-66 year olds, 67+ year olds) i.e. 67+ & 1=67+ year old's houshold includes one other generation, 67+ & 2=67+ year old's houshold includes two other generations. Note 2: Rate per 100 000 person years. Note 3: Stratified by location (UTLA). Note 4: Stratified by location (UTLA) and adjusted for sex, number of comorbidities, categories of housing density (rural or urban setting), smoking status, socio-economic status and includng an interaction between ethnicity and age (as well as the interaction between household composition and ethnicity presented here). Note 5: Stratified on UTLA and adjusted for: sex, smoking, housing density and number of comorbidities and including interactions between ethnicity and: IMD, age and obesity (as well as the interaction with household composition presented here).

**Table S8: Results of statistical tests for interaction between (1) ethnicity and household composition and (2) other variables for which there was evidence of an interaction with ethnicity (for the severe COVID-19 outcome)**

| Variable | p (LRT for interaction) |
|---|---|
| Wave 1 | |
| Household composition | 0.696 |
| Age | <0.0001 |
| Wave 2 | |
| Household composition | 0.003 |
| Age | <0.0001 |
| Deprivation status (IMD) | 0.006 |
| Obesity | <0.0001 |

**Results for the separate severe COVID-19 outcomes (death and hospitalisation)**
Results for analysis of the separate severe COVID-19 outcomes (death and hospitalisation) for wave 1 and wave 2 are provided in Figure S2. Associations between household composition and both of the component outcomes were generally similar to the association between household composition and the (combined) severe COVID-19 outcome for all ethnicities, with the most notable differences for White and South Asian 67+ for the death due to COVID-19 outcome in wave 2. In comparison to the White severe COVID-19 analysis, there was no trend for increasing hazard of the outcome with increasing multigenerational living for COVID-19 death (and all HRs were lower), while for South Asian people there was a steeper trend and larger HRs across the multigenerational categories.

**Results for detailed (16-level) ethnicity analysis for Wave 2**
In analysis of the component ethnicities for the White category (British, Irish, Other White) , British and Other White 67+ year olds were driving the overall effect for Wave 2 (Figure S3), with less evidence of a harmful association for Irish 67+ year olds (although confidence intervals were wide). For the South Asian component ethnicities (Indian, Pakistani, Bangladeshi, Other South Asian), Indian, Pakistani and Other South Asian ethnicities were comparable and consistent with the overall effect, while for Bangladeshi 67+ year olds there was no obvious pattern of increasing harm with increasing generations (although confidence intervals were wide).

**Figure S2. Association between household composition and (1) hospitalisation due to COVID-19 and (2) death due to COVID-19 by ethnicity for wave 1 and wave 2 of the pandemic in England**

Wave 1 models stratified by location (UTLA) and adjusted for sex, number of comorbidities, categories of housing density (rural or urban setting), smoking status, socio-economic status and including an interaction between ethnicity and age (as well as the interaction between household composition and ethnicity presented here). Wave 2 models stratified on UTLA and adjusted for: sex, smoking, housing density and number of comorbidities and including interactions between ethnicity and: IMD, age and obesity (as well as the interaction with household composition presented here).

### Wave 1

#### Admitted to hospital due to COVID-19

| Ethnicity and hh composition | N | Events | HR (95% CI) |
|---|---|---|---|
| **White** | | | |
| Multiple 67+ year olds | 1083552 | 4202 (.4%) | 1.00 (1.00, 1.00) |
| 67+ living alone | 887343 | 6018 (.7%) | 1.35 (1.29, 1.41) |
| 67+ & 1 other gen | 482696 | 2069 (.4%) | 1.14 (1.08, 1.21) |
| 67+ & 2 other gens | 82362 | 372 (.5%) | 1.23 (1.10, 1.37) |
| 67+ & 3 other gens | 12309 | 60 (.5%) | 1.23 (0.95, 1.58) |
| **South Asian** | | | |
| Multiple 67+ year olds | 15264 | 128 (.8%) | 1.00 (1.00, 1.00) |
| 67+ living alone | 14674 | 189 (1.3%) | 1.43 (1.13, 1.80) |
| 67+ & 1 other gen | 26557 | 268 (1%) | 1.18 (0.95, 1.47) |
| 67+ & 2 other gens | 22650 | 263 (1.2%) | 1.34 (1.07, 1.67) |
| 67+ & 3 other gens | 7987 | 101 (1.3%) | 1.35 (1.04, 1.77) |
| **Black** | | | |
| Multiple 67+ year olds | 4019 | 47 (1.2%) | 1.00 (1.00, 1.00) |
| 67+ living alone | 9399 | 143 (1.5%) | 1.20 (0.85, 1.68) |
| 67+ & 1 other gen | 7888 | 86 (1.1%) | 0.95 (0.66, 1.38) |
| 67+ & 2 other gens | 4017 | 46 (1.1%) | 1.22 (0.80, 1.85) |
| 67+ & 3 other gens | 1155 | 13 (1.1%) | 1.23 (0.66, 2.28) |
| **Mixed** | | | |
| Multiple 67+ year olds | 2027 | 13 (.6%) | 1.00 (1.00, 1.00) |
| 67+ living alone | 3129 | 32 (1%) | 1.36 (0.67, 2.77) |
| 67+ & 1 other gen | 2711 | 19 (.7%) | 1.09 (0.51, 2.32) |
| 67+ & 2 other gens | 1209 | 8 (.7%) | 1.12 (0.44, 2.82) |
| **Other** | | | |
| Multiple 67+ year olds | 4581 | REDACT (REDACT%) | 1.00 (1.00, 1.00) |
| 67+ living alone | 6125 | 48 (.8%) | 1.63 (0.93, 2.88) |
| 67+ & 1 other gen | 6185 | 42 (.7%) | 1.68 (0.94, 2.98) |
| 67+ & 2 other gens | 3295 | 20 (.6%) | 1.65 (0.85, 3.19) |
| 67+ & 3 other gens | 771 | REDACT (REDACT%) | 0.94 (0.27, 3.25) |

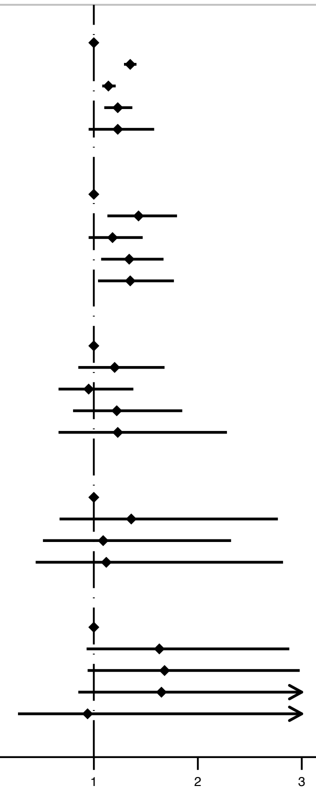


#### Death due to COVID-19

| Ethnicity and hh composition | N | Events | HR (95% CI) |
|---|---|---|---|
| **White** | | | |
| Multiple 67+ year olds | 1083552 | 1972 (.2%) | 1.00 (1.00, 1.00) |
| 67+ living alone | 887343 | 2877 (.3%) | 1.31 (1.23, 1.39) |
| 67+ & 1 other gen | 482696 | 861 (.2%) | 1.09 (1.01, 1.19) |
| 67+ & 2 other gens | 82362 | 140 (.2%) | 1.08 (0.91, 1.29) |
| 67+ & 3 other gens | 12309 | 21 (.2%) | 1.00 (0.65, 1.54) |
| **South Asian** | | | |
| Multiple 67+ year olds | 15264 | 62 (.4%) | 1.00 (1.00, 1.00) |
| 67+ living alone | 14674 | 100 (.7%) | 1.62 (1.17, 2.24) |
| 67+ & 1 other gen | 26557 | 129 (.5%) | 1.26 (0.92, 1.71) |
| 67+ & 2 other gens | 22650 | 138 (.6%) | 1.54 (1.13, 2.09) |
| 67+ & 3 other gens | 7987 | 48 (.6%) | 1.41 (0.96, 2.07) |
| **Black** | | | |
| Multiple 67+ year olds | 4019 | 23 (.6%) | 1.00 (1.00, 1.00) |
| 67+ living alone | 9399 | 67 (.7%) | 1.15 (0.72, 1.85) |
| 67+ & 1 other gen | 7888 | 37 (.5%) | 0.85 (0.51, 1.43) |
| 67+ & 2 other gens | 4017 | REDACT (-%) | 1.15 (0.64, 2.08) |
| 67+ & 3 other gens | 1155 | REDACT (-%) | 0.40 (0.10, 1.70) |
| **Mixed** | | | |
| Multiple 67+ year olds | 2027 | REDACT (-%) | 1.00 (1.00, 1.00) |
| 67+ living alone | 3129 | 12 (.4%) | 0.72 (0.30, 1.75) |
| 67+ & 1 other gen | 2711 | 12 (.4%) | 0.99 (0.41, 2.39) |
| 67+ & 2 other gens | 1209 | REDACT (-%) | 0.60 (0.16, 2.30) |
| **Other** | | | |
| Multiple 67+ year olds | 4581 | 9 (.2%) | 1.00 (1.00, 1.00) |
| 67+ living alone | 6125 | 25 (.4%) | 1.66 (0.72, 3.83) |
| 67+ & 1 other gen | 6185 | 13 (.2%) | 1.07 (0.43, 2.68) |
| 67+ & 2 other gens | 3295 | 11 (.3%) | 1.88 (0.72, 4.90) |

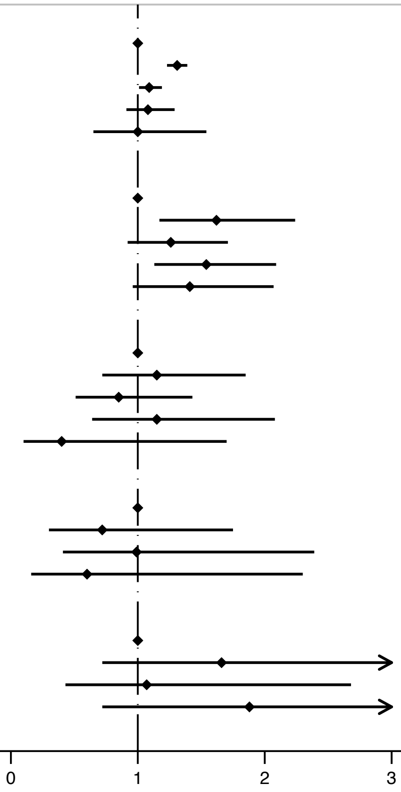

### Wave 2

#### Admitted to hospital due to COVID-19

| Ethnicity and hh composition | N | Events | HR (95% CI) |
|---|---|---|---|
| **White** | | | |
| Multiple 67+ | 1116533 | 9466 (.8%) | 1.00 (1.00, 1.00) |
| 67+ living alone | 880434 | 13414 (1.5%) | 1.37 (1.33, 1.41) |
| 67+ & 1 other gen | 489303 | 4738 (1%) | 1.22 (1.18, 1.27) |
| 67+ & 2 other gens | 84368 | 817 (1%) | 1.27 (1.18, 1.37) |
| 67+ & 3 other gens | 12756 | 169 (1.3%) | 1.64 (1.40, 1.91) |
| **South Asian** | | | |
| Multiple 67+ | 15969 | 268 (1.7%) | 1.00 (1.00, 1.00) |
| 67+ living alone | 14745 | 319 (2.2%) | 1.18 (1.00, 1.40) |
| 67+ & 1 other gen | 27605 | 546 (2%) | 1.17 (1.01, 1.37) |
| 67+ & 2 other gens | 23136 | 571 (2.5%) | 1.42 (1.21, 1.65) |
| 67+ & 3 other gens | 8198 | 273 (3.3%) | 1.74 (1.46, 2.09) |
| **Black** | | | |
| Multiple 67+ | 4132 | 81 (2%) | 1.00 (1.00, 1.00) |
| 67+ living alone | 9462 | 218 (2.3%) | 1.10 (0.84, 1.45) |
| 67+ & 1 other gen | 8050 | 133 (1.7%) | 0.89 (0.67, 1.19) |
| 67+ & 2 other gens | 4119 | 56 (1.4%) | 0.86 (0.61, 1.23) |
| 67+ & 3 other gens | 1185 | 19 (1.6%) | 1.07 (0.64, 1.78) |
| **Mixed** | | | |
| Multiple 67+ | 2117 | 30 (1.4%) | 1.00 (1.00, 1.00) |
| 67+ living alone | 3188 | 52 (1.6%) | 0.97 (0.60, 1.56) |
| 67+ & 1 other gen | 2832 | 32 (1.1%) | 0.78 (0.47, 1.30) |
| 67+ & 2 other gens | 1234 | 20 (1.6%) | 1.15 (0.65, 2.05) |
| **Other** | | | |
| Multiple 67+ | 4807 | 67 (1.4%) | 1.00 (1.00, 1.00) |
| 67+ living alone | 6210 | 102 (1.6%) | 1.04 (0.75, 1.44) |
| 67+ & 1 other gen | 6486 | 75 (1.2%) | 0.83 (0.59, 1.18) |
| 67+ & 2 other gens | 3421 | 43 (1.3%) | 0.99 (0.67, 1.47) |
| 67+ & 3 other gens | 812 | 12 (1.5%) | 1.05 (0.54, 2.05) |

#### Death due to COVID-19

| Ethnicity and hh composition | N | Events | HR (95% CI) |
|---|---|---|---|
| **White** | | | |
| Multiple 67+ | 1116533 | 3371 (.3%) | 1.00 (1.00, 1.00) |
| 67+ living alone | 880434 | 4654 (.5%) | 1.28 (1.22, 1.34) |
| 67+ & 1 other gen | 489303 | 1506 (.3%) | 1.18 (1.11, 1.26) |
| 67+ & 2 other gens | 84368 | 220 (.3%) | 1.06 (0.92, 1.22) |
| 67+ & 3 other gens | 12756 | 46 (.4%) | 1.37 (1.02, 1.83) |
| **South Asian** | | | |
| Multiple 67+ | 15969 | 93 (.6%) | 1.00 (1.00, 1.00) |
| 67+ living alone | 14745 | 118 (.8%) | 1.29 (0.97, 1.70) |
| 67+ & 1 other gen | 27605 | 211 (.8%) | 1.37 (1.07, 1.77) |
| 67+ & 2 other gens | 23136 | 207 (.9%) | 1.53 (1.19, 1.97) |
| 67+ & 3 other gens | 8198 | 115 (1.4%) | 2.12 (1.60, 2.82) |
| **Black** | | | |
| Multiple 67+ | 4132 | 36 (.9%) | 1.00 (1.00, 1.00) |
| 67+ living alone | 9462 | 74 (.8%) | 0.92 (0.61, 1.40) |
| 67+ & 1 other gen | 8050 | 44 (.5%) | 0.74 (0.47, 1.16) |
| 67+ & 2 other gens | 4119 | 21 (.5%) | 0.85 (0.49, 1.48) |
| 67+ & 3 other gens | 1185 | 11 (.9%) | 1.63 (0.82, 3.23) |
| **Mixed** | | | |
| Multiple 67+ | 2117 | 14 (.7%) | 1.00 (1.00, 1.00) |
| 67+ living alone | 3188 | 24 (.8%) | 0.97 (0.49, 1.92) |
| 67+ & 1 other gen | 2832 | 10 (.4%) | 0.56 (0.25, 1.26) |
| 67+ & 2 other gens | 1234 | REDACT (0%) | 1.35 (0.60, 3.05) |
| **Other** | | | |
| Multiple 67+ | 4807 | 21 (.4%) | 1.00 (1.00, 1.00) |
| 67+ living alone | 6210 | 40 (.6%) | 1.40 (0.81, 2.44) |
| 67+ & 1 other gen | 6486 | 15 (.2%) | 0.62 (0.32, 1.22) |
| 67+ & 2 other gens | 3421 | REDACT (0%) | 0.70 (0.30, 1.60) |
| 67+ & 3 other gens | 812 | REDACT (0%) | 1.79 (0.66, 4.89) |

**Figure S3. Association between household composition and severe COVID-19 (death or hospitalisation due to COVID-19) by the component White and South Asian ethnicity categories for wave 2 of the pandemic in England**

Model stratified on UTLA and adjusted for: sex, smoking, housing density and number of comorbidities and including interactions between ethnicity and: IMD, age and obesity (as well as the interaction with household composition presented here).

#### Wave 2: Death or hospital admission due to COVID-19

| Ethnicity and hh composition | Denominator | Events | HR (95% CI) |
|---|---|---|---|
| **British** | | | |
| Multiple 67+ year olds | 1209019 | 10556 (.9%) | 1.00 (1.00, 1.00) |
| 67+ living alone | 831314 | 13325 (1.6%) | 1.36 (1.32, 1.40) |
| 67+ & 1 other gen | 326939 | 3546 (1.1%) | 1.25 (1.20, 1.30) |
| 67+ & 2 other gens | 63308 | 666 (1.1%) | 1.27 (1.17, 1.38) |
| 67+ & 3 other gens | 9562 | 130 (1.4%) | 1.57 (1.32, 1.87) |
| **Irish** | | | |
| Multiple 67+ year olds | 8758 | 96 (1.1%) | 1.00 (1.00, 1.00) |
| 67+ living alone | 8971 | 158 (1.8%) | 1.28 (0.98, 1.65) |
| 67+ & 1 other gen | 4616 | 49 (1.1%) | 0.90 (0.64, 1.28) |
| 67+ & 2 other gens | REDACT | REDACT (REDACT%) | 0.67 (0.31, 1.43) |
| 67+ & 3 other gens | REDACT | REDACT (REDACT%) | 1.93 (0.71, 5.21) |
| **Other White** | | | |
| Multiple 67+ year olds | 52091 | 497 (1%) | 1.00 (1.00, 1.00) |
| 67+ living alone | 43321 | 675 (1.6%) | 1.29 (1.14, 1.45) |
| 67+ & 1 other gen | 21818 | 268 (1.2%) | 1.30 (1.11, 1.51) |
| 67+ & 2 other gens | 9604 | 99 (1%) | 1.33 (1.06, 1.66) |
| 67+ & 3 other gens | 2463 | 28 (1.1%) | 1.44 (0.98, 2.10) |
| **Indian** | | | |
| Multiple 67+ year olds | 13119 | 207 (1.6%) | 1.00 (1.00, 1.00) |
| 67+ living alone | 8059 | 181 (2.2%) | 1.29 (1.05, 1.59) |
| 67+ & 1 other gen | 14151 | 249 (1.8%) | 1.14 (0.94, 1.38) |
| 67+ & 2 other gens | 10807 | 230 (2.1%) | 1.40 (1.15, 1.70) |
| 67+ & 3 other gens | 2844 | 67 (2.4%) | 1.51 (1.13, 2.02) |
| **Pakistani** | | | |
| Multiple 67+ year olds | 2284 | 74 (3.2%) | 1.00 (1.00, 1.00) |
| 67+ living alone | 2576 | 78 (3%) | 0.92 (0.67, 1.28) |
| 67+ & 1 other gen | 5268 | 165 (3.1%) | 1.01 (0.76, 1.34) |
| 67+ & 2 other gens | 6907 | 235 (3.4%) | 1.11 (0.85, 1.46) |
| 67+ & 3 other gens | 3566 | 168 (4.7%) | 1.49 (1.12, 1.98) |
| **Bangladeshi** | | | |
| Multiple 67+ year olds | 256 | 12 (4.7%) | 1.00 (1.00, 1.00) |
| 67+ living alone | 440 | 14 (3.2%) | 0.64 (0.27, 1.53) |
| 67+ & 1 other gen | 760 | 23 (3%) | 0.65 (0.30, 1.43) |
| 67+ & 2 other gens | 1128 | 57 (5.1%) | 1.07 (0.52, 2.20) |
| 67+ & 3 other gens | 667 | 20 (3%) | 0.61 (0.27, 1.36) |
| **Other South Asian** | | | |
| Multiple 67+ year olds | 4170 | 62 (1.5%) | 1.00 (1.00, 1.00) |
| 67+ living alone | 3691 | 68 (1.8%) | 1.17 (0.82, 1.67) |
| 67+ & 1 other gen | 4542 | 90 (2%) | 1.28 (0.92, 1.80) |
| 67+ & 2 other gens | 3500 | 74 (2.1%) | 1.41 (0.99, 2.00) |
| 67+ & 3 other gens | 1060 | 25 (2.4%) | 1.42 (0.88, 2.28) |

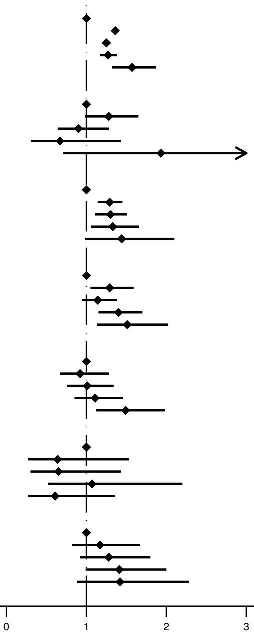

**Analysis of the impact of household size**

Cross-tabulation of household composition against household size (Table S9 - panel 2) showed that increasing household size within categories of household composition (and vice versa) did not correspond to an increasing hazard of severe COVID-19 for either White or South Asian older people (apart from a trend in increasing hazards with increasing household size within the White 67+ & 1 generation). Of note was that for White 67+, across the combined household composition-household size exposure variable, two of the three highest HRs were within the largest household size category (5+) and for South Asian over 67+ year olds all three of the highest HRs were within the largest household size category.

**Table S9: Analysis of how the association between household compostion and severe COVID-19 varies by household size (number of occupants) via cross tabulation of results from a combined household size – household composition variable for White and South Asian ethnicities for Wave 2**

*Wave 2 (1st September – 31st January 2021)*

**Combined household composition/household size exposure variable - multivariable adjusted HR[1] (95% CI)** | **Cross-tabulation of multivariable adjusted results for the household composition/household size exposure variable (household composition rows, household size columns)**

**White**

| Combined household composition/household size exposure variable | Multivariable adjusted HR[1] (95% CI) |
|---|---|
| Two 67+ year olds (hhsize[2]=2) | 1 |
| >2 67+ year olds (hhsize=3-4) | 1·52 (1·30-1·78) |
| 67+ & 1 gen (hhsize=2) | 1·19 (1·14-1·25) |
| 67+ & 1 gen (hhsize=3-4) | 1·23 (1·17-1·29) |
| 67+ & 1 gen (hhsize=5+) | 2·10 (1·74-2·54) |
| 67+ & 2 gen (hhsize=3-4) | 1·27 (1·16-1·39) |
| 67+ & 2 gen (hhsize=5+) | 1·27 (1·13-1·42) |
| 67+ & 3 gen (hhsize=3-4) | 1·54 (1·03-2·30) |
| 67+ & 3 gen (hhsize=5+) | 1·62 (1·38-1·91) |

| | **2** | **3-4** | **5+** |
|---|---|---|---|
| **67+ only** | 1 | 1·49 (1·27-1·75) | |
| **67+ & 1 gen[3]** | 1·18 (1·13-1·24) | 1·23 (1·17-1·29) | 2·08 (1·72-2·52) |
| **67+ & 2 gen** | | 1·25 (1·14-1·37) | 1·24 (1·10-1·40) |
| **67+ & 3 gen** | | 1·44 (0·94-2·19) | 1·57 (1·33-1·85) |

**South Asian**

| Combined household composition/household size exposure variable | Multivariable adjusted HR[1] (95% CI) |
|---|---|
| Two 67+ year olds (hhsize=2) | 1 |
| >2 67+ year olds (hhsize=3-4) | 0·77 (0·41-1·46) |
| 67+ & 1 gen (hhsize=2) | 1·21 (1·00-1·47) |
| 67+ & 1 gen (hhsize=3-4) | 1·15 (0·97-1·37) |
| 67+ & 1 gen (hhsize=5+) | 1·44 (1·08-1·92) |
| 67+ & 2 gen (hhsize=3-4) | 1·03 (0·82-1·29) |
| 67+ & 2 gen (hhsize=5+) | 1·56 (1·33-1·83) |
| 67+ & 3 gen (hhsize=3-4) | 1·34 (0·66-2·72) |
| 67+ & 3 gen (hhsize=5+) | 1·72 (1·44-2·06) |

| | **2** | **3-4** | **5+** |
|---|---|---|---|
| **67+ only** | 1 | 0·77 (0·41-1·46) | |
| **67+ & 1 gen** | 1·21 (1·00-1·47) | 1·15 (0·97-1·37) | 1·44 (1·08-1·92) |
| **67+ & 2 gen** | | 1·03 (0·82-1·29) | 1·56 (1·33-1·83) |
| **67+ & 3 gen** | | 1·34 (0·66-2·72) | 1·72 (1·44-2·06) |

Note 1: Stratified on UTLA and adjusted for: sex, smoking, housing density and number of comorbidities and including interactions between ethnicity and: IMD, age and obesity (as well as the interaction between ethnicity and the combined household composition/size variable presented here). Note 2: Household size (number of occupants in the house). Note 3: p (test-for-trend)=0·003.

**Table S10: Testing the proportional hazards assumption for severe COVID-19 (multivariable analysis)**

| Wave | p-value |
|---|---|
| 1 | 0.596 |
| 2 | 0.467 |

**Table S11: Multivariable-adjusted association between deprivation and (1) severe COVID (hospitalisation or death as a combined outcome) and (2) death from a cause other than COVID-19 by ethnicity in 67+ year olds for Wave 2**

| | Wave 2 (1st September – 31st January 2021) | | | | |
|---|---|---|---|---|---|
| | **Multivariable HR[1] (95% CI)** | | | | |
| | **White** | **South Asian** | **Black** | **Mixed** | **Other** |
| **Severe COVID-19** | | | | | |
| IMD (p test for interaction=0·006[2]) | | | | | |
| 1 (affluent) | 1 | 1 | 1 | 1 | 1 |
| 2 | 1·13 (1·09-1·17) | 1·53 (1·21-1·94) | 1·80 (1·09-2·99) | 1·00 (0·52-1·95) | 1·05 (0·69-1·58) |
| 3 | 1·25 (1·21-1·30) | 1·67 (1·34-2·08) | 1·73 (1·06-2·80) | 1·30 (0·71-2·38) | 1·09 (0·73-1·63) |
| 4 | 1·39 (1·34-1·45) | 2·00 (1·63-2·47) | 1·48 (0·92-2·39) | 1·30 (0·71-2·39) | 1·17 (0·79-1·73) |
| 5 (deprived) | 1·65 (1·58-1·72) | 2·46 (2·00-3·03) | 1·76 (1·10-2·80) | 2·07 (1·15-3·73) | 1·30 (0·88-1·93) |
| **Death from a cause other than COVID-19** | | | | | |
| IMD (p test for interaction=0·610[2]) | | | | | |
| 1 (affluent) | 1 | 1 | 1 | 1 | 1 |
| 2 | 1·07 (1·03-1·11) | 1·09 (0·81-1·47) | 0·78 (0·43-1·42) | 0·56 (0·24-1·30) | 1·14 (0·67-1·93) |
| 3 | 1·15 (1·11-1·19) | 1·24 (0·95-1·63) | 0·96 (0·56-1·63) | 1·12 (0·57-2·19) | 0·96 (0·56-1·64) |
| 4 | 1·22 (1·18-1·27) | 1·28 (0·99-1·66) | 0·84 (0·51-1·41) | 1·16 (0·60-2·23) | 1·17 (0·71-1·94) |
| 5 (deprived) | 1·34 (1·29-1·40) | 1·27 (0·97-1·65) | 1·25 (0·77-2·03) | 1·55 (0·83-2·89) | 1·41 (0·85-2·33) |

Note 1: Stratified on UTLA and adjusted for: sex, smoking, housing density and number of comorbidities and including interactions between ethnicity and: household composition, age and obesity (as well as the interaction between ethnicity and IMD presented here). Note 2: LRT test

**Table S12: Results of senstivitiy analyses (for the death or hospitalisation from COVID-19 combined outcome) relating to (1) including a 5 year buffer between the 67+ year old generation and the next youngest generation (2) only including people who lived in households with 100% TPP coverage (3) using multiple imputation to account for missing ethnicity and (4) a complete records analysis for BMI and smoking.**

| Wave 2 (1st September – 31st January 2021) – MV HR (95% CI) | | | | |
|---|---|---|---|---|
| | Sensitivity analyses | | | |
| Main MV adjusted analysis results (for reference) | 5 year buffer | 100% TPP coverage | Missing ethnicity multiply imputed | Complete records BMI and smoking |
| White | | | | |
| 1 | 1 | 1 | 1 | 1 |
| 1·37 (1·33-1·41) | 1·36 (1·32-1·39) | 1·37 (1·33-1·41) | 1·52 (1·48-1·56) | 1·39 (1·33-1·45) |
| 1·22 (1·17-1·26) | 1·24 (1·20-1·29) | 1·25 (1·20-1·30) | 1.21 (1.18-1.25) | 1·29 (1·21-1·36) |
| 1·26 (1·17-1·36) | 1·27 (1·18-1·37) | 1·27 (1·17-1·38) | 1.21 (1.13-1.29) | 1·44 (1·29-1·62) |
| 1·61 (1·38-1·87) | 1·55 (1·33-1·81) | 1·46 (1·23-1·74) | 1.42 (1.23-1.64) | 1·75 (1·37-2·24) |
| South Asian | | | | |
| 1 | 1 | 1 | 1 | 1 |
| 1·22 (1·03-1·44) | 1·17 (1·00-1·36) | 1·17 (0·99-1·38) | 1.38 (1.19-1.61) | 1·15 (0·91-1·44) |
| 1·22 (1·05-1·42) | 1·15 (1·00-1·33) | 1·12 (0·96-1·31) | 1.25 (1.09-1.44) | 1·11 (0·90-1·36) |
| 1·45 (1·25-1·69) | 1·41 (1·23-1·62) | 1·45 (1·25-1·69) | 1.47 (1.28-1.69) | 1·40 (1·14-1·72) |
| 1·76 (1·48-2·10) | 1·66 (1·41-1·96) | 1·69 (1·41-2·03) | 1.76n (1.49-2.06) | 1·72 (1·35-2·18) |
| Black | | | | |
| 1 | 1 | 1 | 1 | 1 |
| 1·07 (0·82-1·39) | 1·12 (0·88-1·44) | 1·16 (0·90-1·51) | 1.27 (1.01-1.60) | 1·21 (0·85-1·72) |
| 0·88 (0·66-1·17) | 0·94 (0·72-1·24) | 0·89 (0·67-1·19) | 0.88 (0.69-1.13) | 0·93 (0·62-1·37) |
| 0·83 (0·59-1·17) | 0·84 (0·60-1·18) | 0·86 (0·60-1·25) | 0.74 (0.54-1.02) | 0·84 (0·50-1·41) |
| 1·11 (0·69-1·81) | 1·14 (0·70-1·84) | 1·11 (0·63-1·96) | 1.06 (0.68-1.65) | 1·29 (0·60-2·76) |
| Mixed | | | | |
| 1 | 1 | 1 | 1 | 1 |
| 0·90 (0·57-1·42) | 1·07 (0·69-1·66) | 1·21 (0·75-1·93) | 0.74 (0.52-1.05) | 1·10 (0·56-2·17) |
| 0·70 (0·43-1·15) | 0·94 (0·58-1·53) | 0·80 (0·46-1·41) | 0.69 (0.48-1.00) | 0·84 (0·39-1·79) |
| 1·12 (0·65-1·92) | 1·35 (0·79-2·30) | 1·21 (0·64-2·27) | 1.04 (0.69-1.58) | 1·67 (0·74-3·77) |
| 1·62 (0·77-3·41) | 1·71 (0·79-3·67) | 1·31 (0·46-3·73) | 1.20 (0.64-2.24) | 2·56 (0·85-7·65) |
| Other | | | | |
| 1 | 1 | 1 | 1 | 1 |
| 1·10 (0·80-1·51) | 1·15 (0·85-1·55) | 1·22 (0·88-1·68) | 0.90 (0.69-1.17) | 1·05 (0·68-1·63) |
| 0·84 (0·60-1·19) | 0·88 (0·63-1·23) | 0·92 (0·64-1·33) | 0.66 (0.49-0.88) | 0·98 (0·61-1·58) |
| 0·96 (0·65-1·42) | 0·99 (0·68-1·46) | 1·06 (0·69-1·62) | 0.97 (0.71-1.32) | 1·16 (0·66-2·03) |
| 1·12 (0·59-2·13) | 1·18 (0·63-2·22) | 1·33 (0·67-2·61) | 1.32 (0.83-2.10) | 1·05 (0·35-3·15) |

**Table S13: Comparison of OpenSAFELY (TPP) classification of household composition with 2011 Census data**

| | White | | South Asian | | Black | | Mixed | | Other | |
|---|---|---|---|---|---|---|---|---|---|---|
| | Census | TPP | Census | TPP | Census | TPP | Census | TPP | Census | TPP |
| Only 67+ (alone or with other 67+) | 78% | 77% | 33% | 34% | 57% | 51% | 69% | 55% | 47% | 51% |
| 67+ with 1 generation | 19% | 19% | 26% | 31% | 29% | 30% | 23% | 29% | 29% | 29% |
| 67+ with 2 generation | 2% | 3% | 28% | 26% | 12% | 15% | 6% | 19% | 19% | 16% |
| 67+ with 3 generation | 0% | 1% | 13% | 9% | 3% | 4% | 1% | 6% | 6% | 4% |

**References for supplementary material**

1. BETA -Data Security Standards - NHS Digital. NHS Digital. https://digital.nhs.uk/about-nhs-digital/our-work/nhs-digital-data-and-technology-standards/framework/beta---data-security-standards.
2. Data Security and Protection Toolkit - NHS Digital. NHS Digital. https://digital.nhs.uk/data-and-information/looking-after-information/data-security-and-information-governance/data-security-and-protection-toolkit.
3. ISB1523: Anonymisation Standard for Publishing Health and Social Care Data - NHS Digital. NHS Digital. https://digital.nhs.uk/data-and-information/information-standards/information-standards-and-data-collections-including-extractions/publications-and-notifications/standards-and-collections/isb1523-anonymisation-standard-for-publishing-health-and-social-care-data (accessed 11/07/2021).
4. Secretary of State for Health and Social Care - UK Government. Coronavirus (COVID-19): notification to organisations to share information. 2020. https://web.archive.org/web/20200421171727/https://www.gov.uk/government/publications/coronavirus-covid-19-notification-of-data-controllers-to-share-information (accessed 11/07/2021).
5. Williamson, EJ, Walker AJ, Bhaskaran K, *et al.* OpenSAFELY: factors associated with COVID-19 death in 17 million patients. *Nature 2020*; published online July 8 2020. DOI:10.1038/s41586-020-2521-4
6. Ministry of Housing, Communities & Local Government. English indices of deprivation 2015: Index of Multiple Deprivation. Sept 30, 2015. https://www.gov.uk/government/statistics/english- indices-of-deprivation-2015 (accessed August 2, 2021).
